# Natural Progression of Routine Laboratory Markers following Spinal Trauma: A Longitudinal, Multi-Cohort Study

**DOI:** 10.1101/2021.01.19.21250027

**Authors:** Lucie Bourguignon, Anh Khoa Vo, Bobo Tong, Fred Geisler, Orpheus Mach, Doris Maier, John L.K. Kramer, Lukas Grassner, Catherine R. Jutzeler

## Abstract

**Objective:** To track and quantify the natural course of hematological markers over the first year following spinal cord injury.

**Methods:** Data on hematological markers, demographics, and injury characteristics were extracted from medical records of a clinical trial (Sygen) and an ongoing observational cohort study (Murnau Study). The primary outcomes were concentration/levels/amount of commonly collected hematological markers at multiple time-points. Two-way ANOVA and mixed-effects regression techniques were used to account for the longitudinal data and adjust for potential confounders. Trajectories of hematological markers contained in both data sources were compared using the slope of progression.

**Results:** At baseline (≤ 2 weeks post-injury), most hematological markers were at pathological levels, but returned to normal values over the course of six to twelve months post-injury. The baseline levels and longitudinal trajectories were dependent on injury severity. More complete injuries were associated with more pathological values (e.g. hematocrit, ANOVA test; Chisq = 77.10, df = 3, adjusted p-value<0.001, and Chisq = 94.67, df = 3, adjusted p-value<0.001, in the Sygen and Murnau studies, respectively). Comparing the two databases revealed some differences in the hematological markers, which are likely attributable to differences in study design, sample size, and standard of care.

**Conclusions:** Due to trauma-induced physiological perturbations, hematological markers undergo marked changes over the course of recovery, from initial pathological levels that normalize within a year. The findings from this study are important as they provide a benchmark for clinical decision making and prospective clinical trials. All results can be interactively explored on the Haemosurveillance website (https://jutzelec.shinyapps.io/Haemosurveillance/).

**Code availability:** https://github.com/jutzca/Systemic-effects-of-Spinal-Cord-Injury

## Introduction

Owing to its crucial role in the coordination of bodily functions, damage to the spinal cord can lead to severe dysfunction or failure in single or multiple organs, including heart, kidney, and liver.^1^ As a consequence of altered functions, spinal cord-specific cellular components (e.g., enzymes, proteins) are released to the bloodstream^2^ and their quantity relates to the extent of system damage.^3^ In addition to this quantifiable relationship, their readiness and straightforward collection make these hematological markers ideally suited to evaluate the trauma-induced systemic perturbations. Laboratory tests are routinely conducted in the acute phase of injury to assess the initial magnitude of systemic damage and monitor the bodily functions. However, little is known about how the systemic effects and their respective blood markers progress as a function of time. This paucity of knowledge is even more surprising, considering that these hematological markers have the potential to guide the design (patient stratification) and implementation of clinical trials (safety assessment of trialed drug).^4–6^ To address this knowledge gap, the aim of this study was to determine the natural progression of hematological markers following a spinal cord injury. We hypothesized that, by disruption of normal innervation of vital organs after a traumatic spinal cord injury, there will be time-dependent and injury-specific alterations in hematological markers characterized by an initial pathological change that normalize over time (i.e., reach norm values of healthy able-body people). Lastly, we provide the scientific and medical community with a first-of-its-kind surveillance tool ‘Haemosurveillance’ that aims to generate novel research questions as well as to inform clinical decision making and clinical trial design.

## METHODS

### Study design and data source

To determine the natural progression of hematological and serological markers following spinal cord injury, we performed an observational study of prospectively collected data. Therefore, we analyzed two different data sources, one each from the United States of America (USA) and Germany. The first data source was a prospective phase III, placebo controlled, multi-centre study assessing the efficacy of GM-1 ganglioside therapy in acute traumatic spinal cord injury.^7,8^ Running from 1992 to 1998, the Sygen trial failed to demonstrate a superior treatment effect of GM-1 over placebo treatment. Full design, recruitment and enrollment details of the Sygen trial have been described previously.^9^ A total of 797 patients across the USA were included in the randomization. Within the framework of this US Food and Drug Administration (FDA) regulated trial, detailed information concerning neurological scores and blood chemistry were meticulously collected. The second data source was an observational cohort study conducted at the over-regional level-I trauma center in Murnau, Germany (hereafter referred to as Murnau study). Between 2004 and 2017, 363 patients were enrolled and followed-up on for one-year post-injury. All patients enrolled in the Murnau study received standards of rehabilitation care.

### Ethics approval

The study was performed in accordance with the Declaration of Helsinki. Approval for the secondary analysis of the Sygen trial was received by an institutional ethical standards committee on human experimentation at the University of British Columbia. The original Sygen clinical trial (results published elsewhere) also received ethical approval, but was conducted before clinical trials were required to be registered^8–10^. The data received from the original clinical trial were de-identified. The Murnau study was approved by the Bavarian Medical Chamber [#2018-077].

### Cohort definition: inclusion and exclusion criteria

To be included in our study, patients needed to have blood values at three different time-points as well as information on sex, age, and injury characteristics (i.e., injury severity, injury level, and baseline motor and sensory scores). Baseline was defined as the first 72 hours after injury for the Sygen trial and the first two weeks post-injury for the Murnau study. Patients were excluded if any of these data were missing, sustained a non-traumatic injury (e.g., tumor), or decided to withdraw their data over the course of the study.

### Outcome, predictor, and confounding variables

The primary outcomes were hematological and serological markers with data available for at least 50 patients at each time point. This threshold was chosen to ensure that the model output is interpretable, statistically powerful enough to make inferences, and clinically relevant. Independent variables were timepoints post-injury at which hematological and serological markers were collected. As an FDA-requirement for the Sygen trial, detailed information regarding routine blood chemistry was collected at admission to the trauma-center (hereinafter referred to as week 0), 1, 2, 4, 8, and 52 weeks post-injury. In the Murnau study, information on hematological and serological markers were collected upon request of the attending physicians (i.e., not at standardized time-points). As a consequence, different numbers of blood draws were collected for each patient on different days post-injury. Potential confounders included age, sex, injury completeness (at time of injury) according to the American Spinal Injury Association (ASIA) Impairment Scale (AIS),^11^ and level of injury (at/above T6 vs below T6).

### Statistical Analyses

Two-way analysis of variance (two-way ANOVA) and mixed effects regression models were chosen for the primary analyses. These models naturally suited to account for the longitudinal nature of the data as well as to adjust for potential confounders. Dependent variables were all hematological and serological markers that met our inclusion criteria. In the Murnau study, blood values were averaged per week, from week 0 to week 7 post-injury. In both studies, if, for a certain marker, patient and time point, no data was available, the time point for this patient’s marker was excluded. For analyses comparing both studies, we examined the percentage of deviation from the mean of the normal range, collected from the Murnau study. The rationale for this normalizing procedure was to make the data of the two cohorts comparable despite having different units. Independent variables were time post-injury, AIS grade, or level of injury, when examining data from the individual studies. When comparing the hematological markers from both studies, we added the data sources as an independent variable. For mixed effects regression models, pairwise comparisons of the different levels of the independent variable of interest were performed. Hence, significance levels were adjusted for multiple comparisons using Tukey’s test and p < 0.05, after adjustment, was regarded as statistical significance. For one-study two-way ANOVA tests, we applied Bonferroni correction for testing for six independent variables together. Thus, we adjusted p-values and p<0.05 was regarded as statistical significance. In the same way, when comparing the two studies, no correction was applied as only the data source was considered as an independent variable. Thus, p < 0.05 was regarded as statistical significance. For all analyses, R Statistical Software, version 3.6.3 (running under: macOS Mojave 10.13.6), was used.

### Data visualization

Using the R package *Shiny* and *ShinyDashboard*, we created an online interface to visualize the results of the current study and to interactively explore the data used for this study.

### Data and code availability statement

Anonymized data used in this study will be made available upon request to the corresponding author and in compliance with the General Data Protection Regulation (EU GDPR). The code describing the analysis can be accessed on our GitHub repository (https://github.com/jutzca/Systemic-effects-of-Spinal-Cord-Injury).

## RESULTS

### Cohort summary: Included patients

Subject and injury characteristics of both cohorts (Sygen: 679; Murnau: 239) are summarized in **Table 1**. A comparison revealed a comparable ratio of male and female patients (Pearson’s chi-squared test, X-squared = 0.07, df = 1, p = 0.786). However, significant differences were found in terms of age distribution (two-sided t-test, t = 13.63, df = 322.55, p < 0.001, **Figure e-1**) and injury severity distribution (Pearson’s chi-squared test, X-squared = 244.9, df = 3, p < 0.001).

**Table 1:**
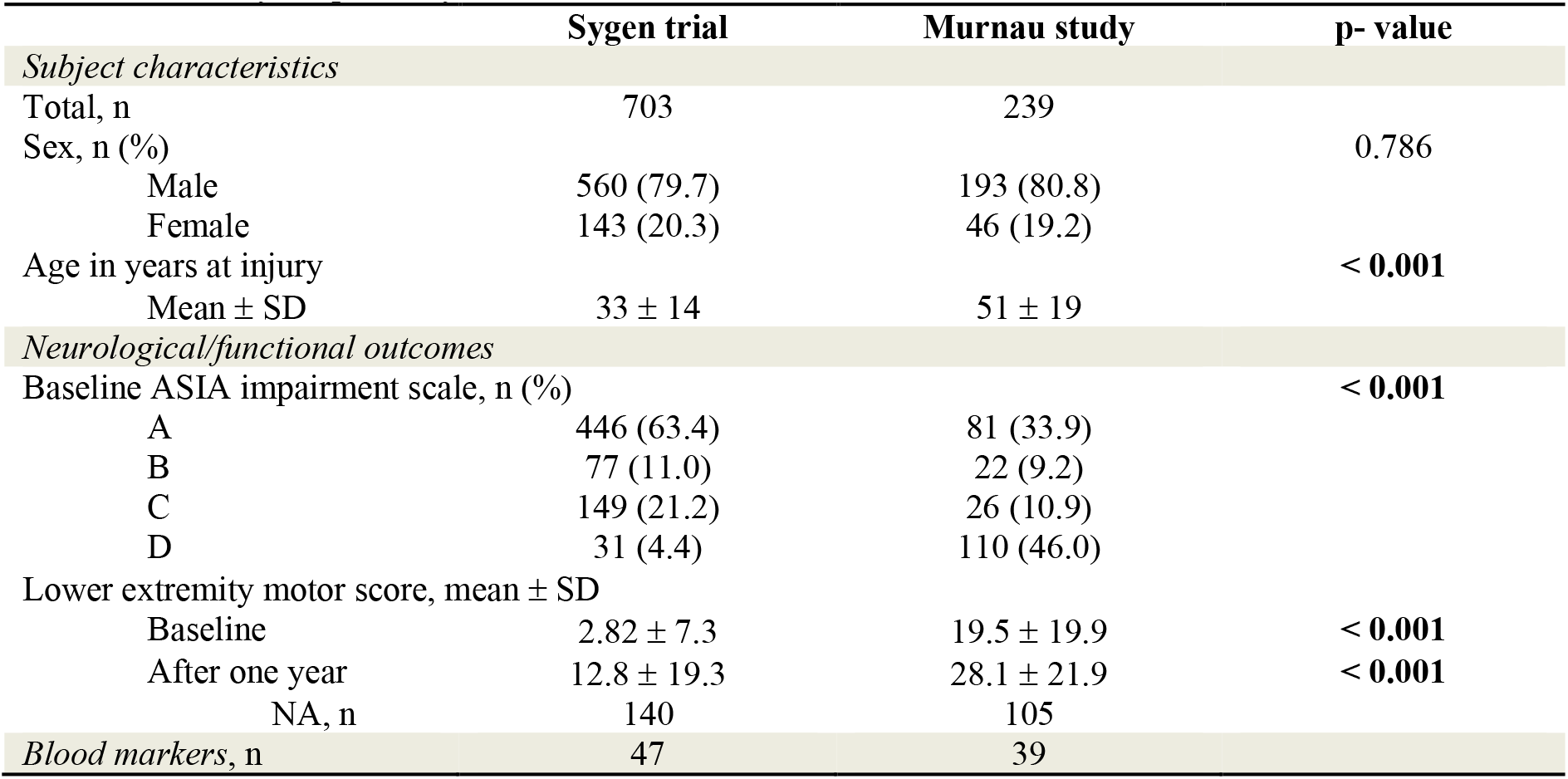
Subject and injury characteristics of patients included in our analysis and enrolled in the Sygen trial and Murnau study, respectively.

|  | Sygen trial | Murnau study | p- value |
| --- | --- | --- | --- |
| *Subject characteristics* |  |  |  |
| Total, n | 703 | 239 |  |
| Sex, n (%) |  |  | 0.786 |
| Male | 560 (79.7) | 193 (80.8) |  |
| Female | 143 (20.3) | 46 (19.2) |  |
| Age in years at injury |  |  | < 0.001 |
| Mean ± SD | 33 ± 14 | 51 ± 19 | |
| *Neurological/functional outcomes* |  |  |  |
| Baseline ASIA impairment scale, n (%) |  |  | < 0.001 |
| A | 446 (63.4) | 81 (33.9) |  |
| B | 77 (11.0) | 22 (9.2) |  |
| C | 149 (21.2) | 26 (10.9) |  |
| D | 31 (4.4) | 110 (46.0) |  |
| Lower extremity motor score, mean ± SD | | | |
| Baseline | 2.82 ± 7.3 | 19.5 ± 19.9 | < 0.001 |
| After one year | 12.8 ± 19.3 | 28.1 ± 21.9 | < 0.001 |
| NA, n | 140 | 105 |  |
| *Blood markers, n* | 47 | 39 |  |

### Cohort summary: Excluded patients

A total of 94 and 124 patients in the Sygen trial and Murnau study, respectively, did not meet the inclusion criteria and were excluded. Reasons for exclusion comprised normal AIS score (AIS E, n = 5) and missing information on baseline AIS score (n = 192). **Table e-1** provides a detailed overview of the excluded cohorts. Excluded and included cohorts were significantly different in terms of age distribution (two-sided t-test; t = 2.03, df = 124.56, p = 0.04, with excluded cohort younger than included cohort; and, t = −1.8852, df = 123.91, p = 0.06, with excluded cohort older than included cohort), in the Sygen trial and Murnau study, respectively. Excluded and included cohorts were comparable in terms of ratio of male and female patients (Pearson’s chi-squared test; X-squared = 3.43, df = 1, p = 0.06), in the Sygen trial, but significantly different in the Murnau study (Pearson’s chi-squared test; X-squared = 8.73, df = 1, p = 0.003).

### Hematological and serological markers

A total of 32 and 28 routinely assessed blood markers were available in the Sygen trial and Murnau study, respectively. Among these, 14 and 8 blood markers, respectively, were part of the complete blood count (CBC), which is a test that evaluates the cells that circulate in blood. Notably, it includes counts of platelets, red and white blood cells, hemoglobin, and hematocrit. The remaining blood markers reflect renal function (5 and 4 markers in the Sygen trial and Murnau study, respectively), hepatic function (5 and 6 markers), pancreatic function (1 and 2 markers) and muscle damages (2 and 3 markers). Overall, 20 blood markers were shared among the two data sources. **Table e-2** provides an overview of all collected markers.

### Natural progression of hematological and serological markers post-injury

With the exception of amylase, gamma-glutamyl transferase (GGT), glucose, lipase and alanine aminotransferase (ALAT) in the Murnau study (p = 0.450, p = 1, p = 0.074, p = 1, p = 0.218, respectively) and alkaline phosphatase, potassium, mean corpuscular volume (MCV) and thrombocyte levels in the Sygen trial (p = 0.554, p = 1, p = 1, p = 1, respectively), the concentrations of hematological markers significantly changed as a function of time since injury (**Table e-3 and Table e-4**). For 28 hematological markers, these changes occurred within the normal range. The remaining 24 hematological markers had baseline values outside the normal range, which normalized over the course of recovery (**Figure 1** and **Figure 2**). One (i.e., hematocrit) blood marker remained outside the normal range at 1-year post-injury.

**Figure 1.**
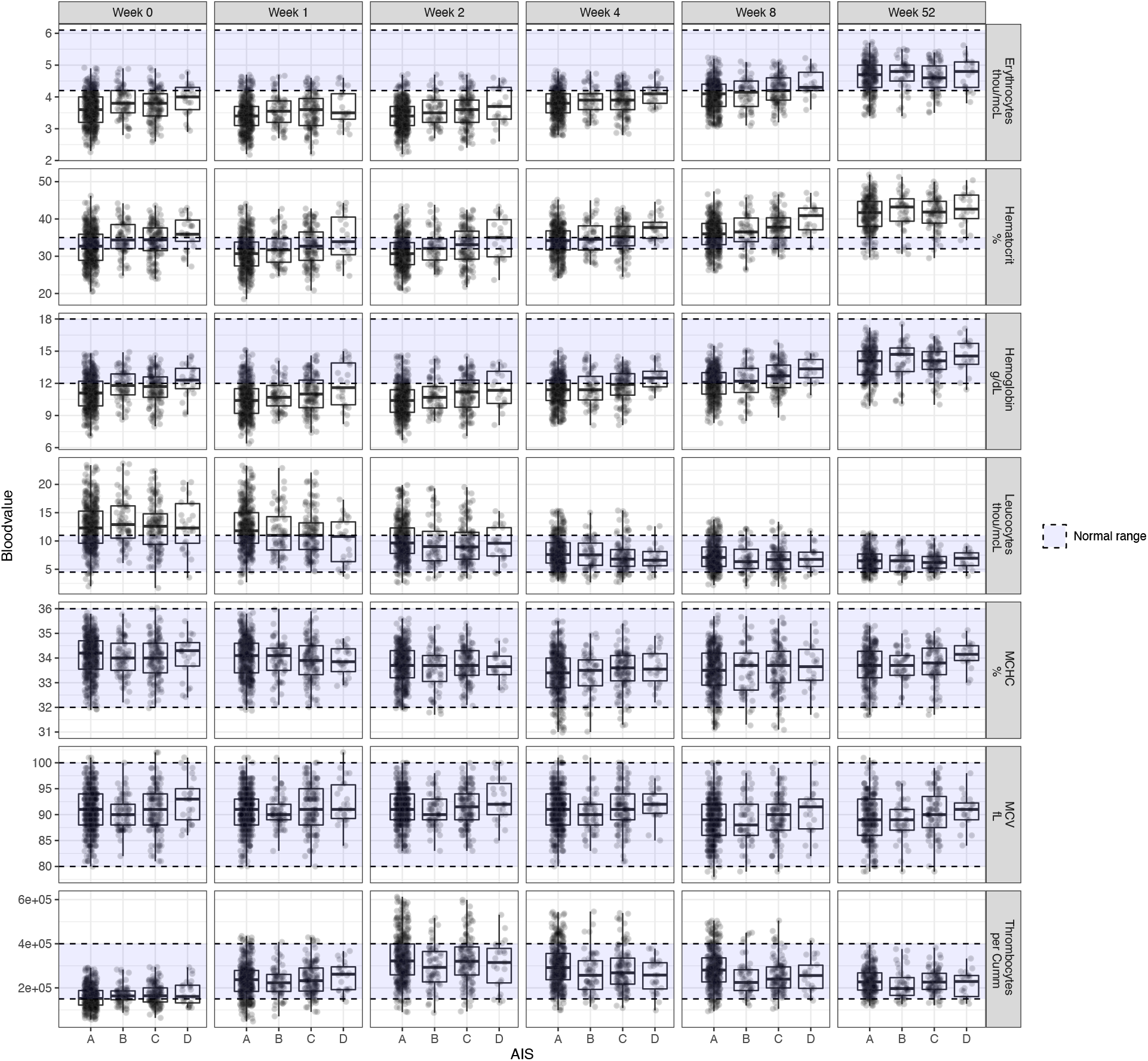
Natural progression of the complete blood count in patients with spinal cord injury that were enrolled in the Sygen trial. Three different patterns of progression were observed. Firstly, the blood markers remained constant and within the range of able-body people, such as mean corpuscular volume (MCV) and mean corpuscular hemoglobin (MCH). Secondly, blood markers are pathological immediately after the trauma, but recover over the course of a year and reach the normal range. Erythrocytes, haemoglobin, and leucocytes are characterized by such a course. Thirdly, values are initially within the normal range, but as a function of time they become pathological when compared to able-bodied people. Hematocrit is one such example. For clinical decision making as well as the design and implementation of clinical trials, it is of utmost importance to know the temporal progression of these blood markers.

**Figure 2.**
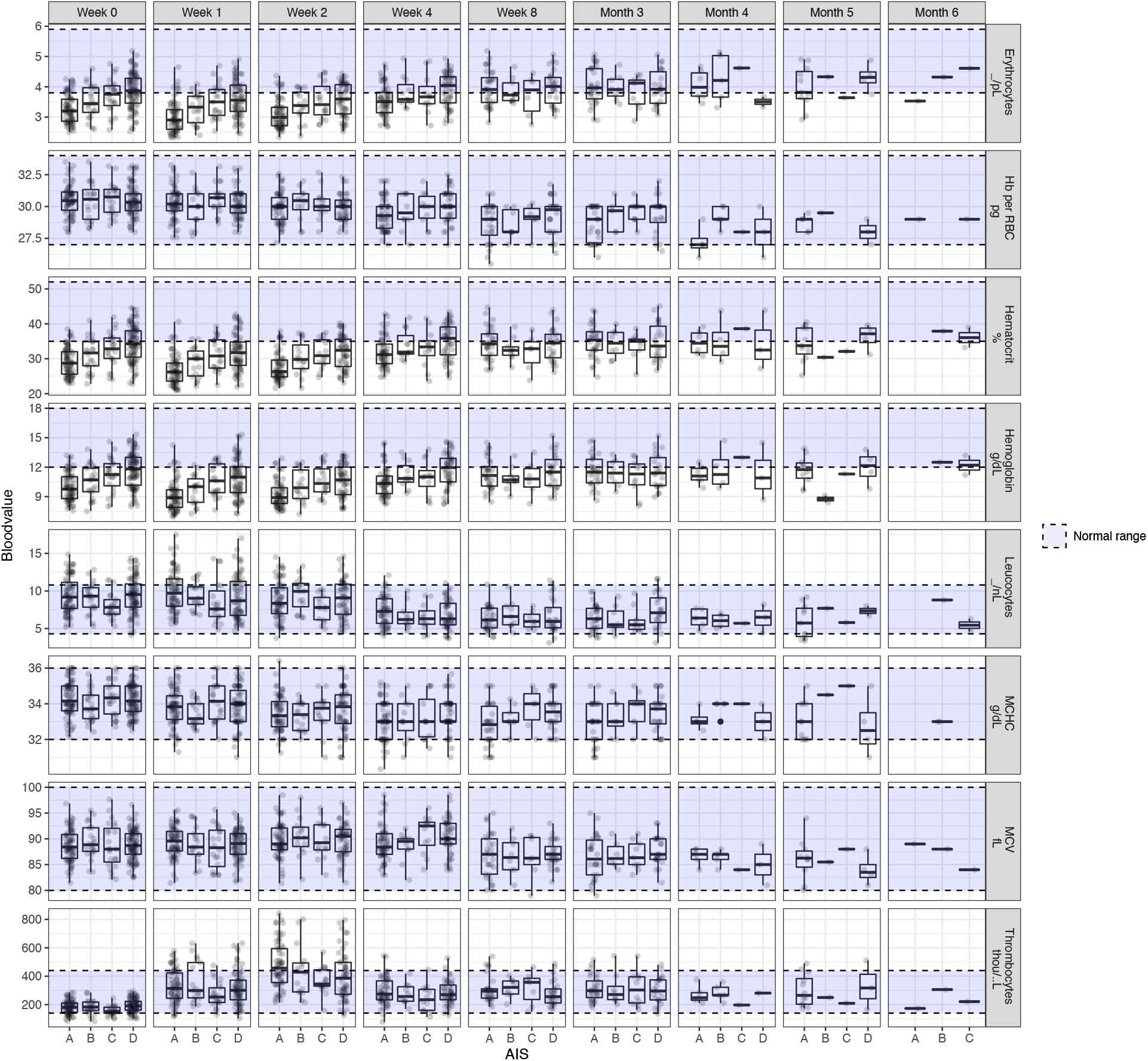
Natural progression of the complete blood count in patients with spinal cord injury that were enrolled in the Murnau study.

### Relationship between hematological levels and injury characteristics

In line with our hypothesis, the ANOVA revealed a global effect of injury severity (i.e., AIS score). Our post-hoc analysis revealed that the hematological values were dependent on the AIS grades, particularly amylase (p=1.94e-2 and p=9.36e-3), calcium (p<0.001 and p<0.001), hematocrit (p<0.001 and p<0.001), hemoglobin (p<0.001 and p<0.001), erythrocytes count (p<0.001 and p<0.001), and total protein/albumin levels (p<0.001 and p<0.001), in both Murnau study and Sygen trial, respectively **(Table e-3 and Table e-4**). The pairwise comparisons between the AIS grades yielded that calcium, hematocrit, hemoglobin, erythrocytes count, and total protein/albumin levels were significantly different between patients classified as AIS A and D. In all cases, higher values for these markers, closer to the normal range, were associated with less severe injury (AIS D), as illustrated in **Figure 1**. Additionally, hematocrit, hemoglobin, erythrocytes count, and total protein/albumin were significantly different between patients classified as AIS A and C, as well as AIS B and D. All results are reported in **Table e-5 and Table e-6** and illustrated in **Figures e-2** to **Figure e-6**. In terms of injury level, we found no significant differences in hematological values between patients with injuries at/above T6 compared to those with injuries below T6 in both the Murnau study and Sygen trial **(Table e-3 and Table e-4)**.

### Comparison between historical and contemporary cohort

As described above, the Murnau study and Sygen trial come with a number of major differences in their design. As illustrated in **Figure 3**, there were significant differences in the hematological markers and their progression (**Table e-7**), with the exception of amylase (p = 0.103), alkaline phosphatase (p = 0.370), mean corpuscular hemoglobin concentration (MCHC, p = 5.23e-2), MCV (p = 0.647), and alanine aminotransferase (ALAT) levels (p = 0.718).

**Figure 3.**
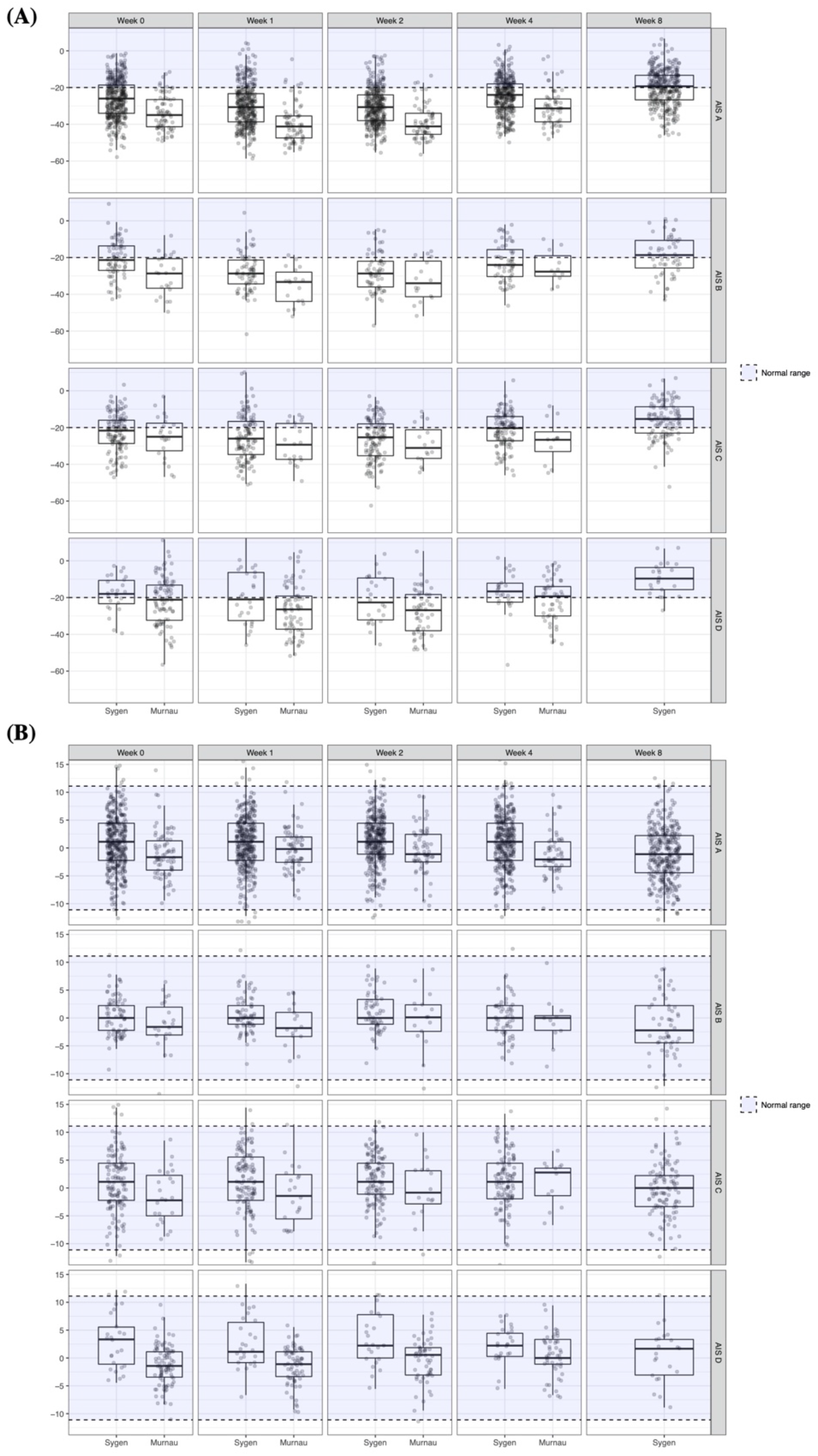
Comparison of the natural progression of hemoglobin (A) and mean corpuscular volume (B) in patients with spinal cord injury enrolled in the Sygen trial and Murnau study, respectively.

### Data visualization

All results can be explored interactively on the Haemosurveillance website (https://jutzelec.shinyapps.io/Haemosurveillance/). Information is presented in separate tabs for patients enrolled in Sygen and Murnau data, respectively. The interactive interface also allows to visualize the data stratified by demographics (sex and age group) and injury characteristics (i.e., injury severity and type of plegia). Additionally, the interface facilitates a direct comparison of the two data sources.

## DISCUSSION

The present study describes the natural progression of hematological parameters that are routinely assessed on admission and in the days to weeks following acute spinal cord injury. Consistent with our first hypothesis, we found trauma-induced changes in routinely collected hematological markers (e.g., hemoglobin, glucose). By in large, most of markers normalized at 1-year post-injury (i.e., reached norm values of healthy able-body people). Our second hypothesis was also confirmed, insofar as the observed changes in markers were dependent on age at injury, sex, and injury severity, but not injury level. This suggests that these changes, in addition to reflecting the polytrauma and the consequent recovery process, are also capturing the severity of the spinal cord injury. Collectively, this study provides new insights that will aid the design and implementation of clinical trials.

### Natural progression and the relationship between hematological levels and injury severity

In the present study, the majority of the hematological markers reach pathological level shortly after the traumatic event and then normalize within a year post-injury. At baseline (within 2 weeks post injury), the degree of alterations in the hematological markers was associated with the injury severity, in such a way that patients with complete injuries exhibited more pronounced abnormalities in hematological markers compared to those with incomplete injuries. This relationship between hematological markers and degree of injury severity underpins the notion that hematological markers may be utilized as measurable indicators of the severity. As such, they can aid the diagnosis of spinal cord injury severity, particularly in cases where standard neurological examination is not possible (e.g., intoxicated or unresponsive patients).^4^ Moreover, abnormalities in certain hematological markers may also induce further damage or delay the recovery process (e.g., albumin)^12,13^ and thus, need to be addressed. In a recent study, Tong and colleagues detected that patients with prolonged hypoalbuminemia recovered to a lesser degree than those patients with normal albumin levels^13^. Timely substitution of albumin might have beneficial effects on the functional and neurological recovery of the patient as suggested by findings from animal studies^14^. While the return to normal hematological levels occurs along the same timeline as the neurological and functional recovery, for many hematological markers there is no longer an association between hematological levels and injury severity. This lack of association in the chronic phase of injury suggests that the hematological markers are more representative of the initial polytrauma and the recovery from it as opposed to be specific indices of the spinal cord injury.

### Hematological markers in the design and implementation of clinical trials

Our study provides an important framework for the implementation of hematological markers in the design and conduction of clinical trials. Conventionally, the safety and tolerability of trialed treatments are assessed by means of specific abnormalities of routinely collected the hematological and CSF markers.^15^ As the majority of drugs, including the currently trialed riluzole^16,17^ and minocycline,^18,19^ are metabolized and cleared by the liver and kidney, respectively, regulatory agencies released guidelines for the assessment of risk surrounding drug-induced liver injuries (DILI)^20,21^ and nephrotoxicity^22^ in clinical trials. Multiple scheduled blood draws facilitate the early detection, tracking, and management of drug-induced organ damage. Typically, any deviation from the norm values of healthy able-body people would alert the investigators. In spinal cord injury, however, baseline values of numerous hematological markers are pathological (**Figures 1** and **2**), which, when ignored or unknown, can substantially bias assumptions on drug safety. Our Haemosurveillance tool offers a first-of-its-kind platform to accurately disentangle drug-induced from trauma-driven perturbations in routinely collected hematological markers. This tool is particularly useful for (1) clinical trials without a control group (i.e., placebo) and (2) clinical trials with a control group that is not being managed by standard of care. In the former, historical data can aid evaluation of safety of the trialed drug, while in the latter the effect of the deviation from the standard of care can be measured. For example, in the ongoing NISCI trial (https://nisci-2020.eu/index.php?id=1449), all enrolled patients are subject to repeated lumbar puncture regardless of their allocation. As repeated lumbar puncture is not standard of care, historical data can be leveraged to assess their impact on health is unknown (e.g., rate of infections).

In addition to providing guidance in drug safety and tolerability, hematological markers bear the potential to refine the stratification of patients and increase the likelihood to detect a significant treatment effect.^23,24^ A major barrier to detecting small treatment effects in clinical trials is the extensive heterogeneity of the neurological recovery and the scarcity of reliable predictors, such as the initial damage to the spinal cord (i.e., AIS scores), that can fully capture the extent of the injury. Thus, utilizing a biological correlate (e.g., blood or CNS marker) is potentially advantageous and informative owing to its representation of the trauma and indirect involvement in the central nervous system.

### Differences between data sources

In the current study, we analyzed data from two different data sources to validate our findings regarding temporal trajectories of the hematological markers. Overall, these trajectories show comparable trends. However, some differences were uncovered that are likely attributable to differences in the study design, study period, standard of care, population structure, and sample size. The Sygen trial, our first data source, was conducted in the ‘90s and had five pre-defined time-points of blood collection. Moreover, as part of the standard of care at the time, all patients sustaining a spinal cord injury received methylprednisolone, a corticosteroid, to reduce inflammation and secondary damage.^25,26^ Corticosteroids have been reported to alter the concentration of certain hematological markers, including bilirubin, albumin, and leukocytes.^27–29^ Patients enrolled in the Sygen trial exhibited reduced bilirubin levels and leukocytosis (i.e., an increase in the number of white cells in the blood) compared to the patients in the Murnau study that did not receive acute treatment of methylprednisolone. Moreover, the time-points of blood draw could have contributed to the differences observed. While the Sygen trial collected blood samples at predefined time points, the patients in the observational Murnau study were subject to blood draws when indicated by the treating physician. Lastly, it is well-known that organ function declines with age and is correlated with changes in laboratory values. A larger proportion of elderly patients was enrolled in the Murnau study and thus, could have contributed to the divergent findings.^30,31^

### Limitations

The primary limitation of the current study is that we utilized retrospective data, from a nearly 20-year-old clinical trial. We partially address this limitation by prospectively collecting contemporary data in the framework of the Murnau study. Potential bias introduced by changes in standards of care over the last decades can be, at least in part, mitigated. However, time-points of data collection was not standardized in the Murnau study. As a consequence, the time-varying sample size complicated the analyses. Future studies with larger and more consistent sample sizes at each time-point of data collection are warranted to validate our findings. The small sample size further prevented a meaningful subgroup analysis stratified by sex and age, considering that many hematological markers have different normal ranges for women and men as well as are subject to age-related changes. It should also be noted that excluding patients due to missing AIS grade (e.g. because the patient was unconscious at baseline) represents a loss of information and introduces a potential bias towards patients with slightly less severe injuries. Studies with large sample sizes at baseline and follow-up timepoints are warranted to address this in further detail. Additionally, our study is focused on correlations at the population level, which does not guarantee the translation of our findings at the individual level. Further investigations are needed to assess the potential of hematological markers in individual recovery prediction. Moreover, we did not account for any of the medications that were administered to the patients to treat secondary complications associated with spinal cord injury.^32,33^ Some medications (e.g., corticosteroids and nonsteroidal anti-inflammatory [NSAID] medication) can affect the concentration of the hematological markers. Future studies should also address the impact of medication on the hematological markers, particularly in the acute phase of injury.

### Conclusion

To our best knowledge, this is the first study to comprehensively investigate the natural progression of hematological markers in patients with a traumatic spinal cord injury. As a consequence of the sustained trauma, numerous routinely collected hematological markers are altered in their concentration. The majority of these makers return to a normal range after 6-12 months post-injury. Importantly, the findings from this study provide a benchmark for clinical decision making and prospective clinical trials. Our online surveillance platform (Haemosurveillance) provides a tool for the spinal cord injury community, researchers, authorities, and policymakers to interactively exploit the natural progression of hematological markers and compare different datasets with each other. The platform is configured such that existing or newly generated datasets can be added if they comply with GDPR.

## Acknowledgments

The authors would like to acknowledge the participating centers in the EMSCI network (http://emsci.org/members) that were involved in the patient care and collection of data necessary for this study.

## Appendix 1: Authors

| **Name** | **Location** | **Contribution** |
| --- | --- | --- |
| Lucie Bourguignon, MSc | ETH Zurich and SIB Swiss Institute of Bioinformatics, Basel, Switzerland | data cleaning, data analyses, interpretation of data, and drafting the manuscript |
| Anh Kho Vo, MSc | University of British Columbia, Vancouver, Canada | data analyses, interpretation of data, and revising the manuscript for intellectual content |
| Bobo Tong, MPH | University of British Columbia, Vancouver, Canada | data cleaning, interpretation of data and revising the manuscript for intellectual content |
| Fred Geisler, MD PhD | University of Saskatchewan, Saskatoon, Saskatchewan, Canada | primary data collection, interpretation of data, and revising the manuscript for intellectual content |
| Orpheus Mach, MA | Spinal Cord Injury Center, Trauma Center Murnau, Murnau, Germany | primary data collection, and revising the manuscript for intellectual content |
| Doris Maier, MD | Spinal Cord Injury Center, Trauma Center Murnau, Murnau, Germany | primary data collection, and revising the manuscript for intellectual content |
| John LK Kramer, PhD | University of British Columbia, Vancouver, Canada | study concept/design, interpretation of data, and revising the manuscript for intellectual content |
| Lukas Grassner, MD PhD | Department of Neurosurgery, Medical University Innsbruck, Innsbruck, Austria | primary data collection, interpretation of data, and revising the manuscript for intellectual content |
| Catherine R. Jutzeler, PhD | ETH Zurich and SIB Swiss Institute of Bioinformatics, Basel, Switzerland | study concept/design, data entry, data cleaning, data analyses, interpretation of data, and drafting the manuscript |

## Notes

### Competing Interest Statement

The authors have declared no competing interest.

### Funding Statement

This study was funded by research grants from the Swiss National Science Foundation (Ambizione Grant #PZ00P3_186101, Jutzeler) and Wings for Life Research Foundation (#2017_044, Jutzeler and Kramer).

### Author Declarations

The study was performed in accordance with the Declaration of Helsinki. Approval for the secondary analysis of the Sygen trial was received by an institutional ethical standards committee on human experimentation at the University of British Columbia. The original Sygen clinical trial (results published elsewhere) also received ethical approval, but was conducted before clinical trials were required to be registered. The data received from the original clinical trial were de-identified. The Murnau study was approved by the Bavarian Medical Chamber [#2018-077].

## References

1. Stein, D.M., Menaker, J., McQuillan, K., Handley, C., Aarabi, B., and Scalea, T.M. (2010). Risk factors for organ dysfunction and failure in patients with acute traumatic cervical spinal cord injury. Neurocrit. Care.

2. Yokobori, S., Zhang, Z., Moghieb, A., Mondello, S., Gajavelli, S., Dietrich, W.D., Bramlett, H., Hayes, R.L., Wang, M., Wang, K.K.W., and Bullock, M.R. (2015). Acute diagnostic biomarkers for spinal cord injury: Review of the literature and preliminary research report. World Neurosurg. 83, 867–878.

3. Kwon, B.K., Stammers, A.M.T., Belanger, L.M., Bernardo, A., Chan, D., Bishop, C.M., Slobogean, G.P., Zhang, H., Umedaly, H., Giffin, M., Street, J., Boyd, M.C., Paquette, S.J., Fisher, C.G., and Dvorak, M.F. (2010). Cerebrospinal fluid inflammatory cytokines and biomarkers of injury severity in acute human spinal cord injury. J. Neurotrauma.

4. Kwon, B.K., Bloom, O., Wanner, I.B., Curt, A., Schwab, J.M., Fawcett, J., and Wang, K.K. (2019). Neurochemical biomarkers in spinal cord injury. Spinal Cord.

5. Kwon, B.K., Streijger, F., Fallah, N., Noonan, V.K., Bélanger, L.M., Ritchie, L., Paquette, S.J., Ailon, T., Boyd, M.C., Street, J., Fisher, C.G., and Dvorak, M.F. (2016). Cerebrospinal Fluid Biomarkers To Stratify Injury Severity and Predict Outcome in Human Traumatic Spinal Cord Injury. J. Neurotrauma, neu.2016.4435.

6. Badhiwala, J.H., Wilson, J.R., Kwon, B.K., Casha, S., and Fehlings, M.G. (2018). A Review of Clinical Trials in Spinal Cord Injury including Biomarkers. J. Neurotrauma 1917, eu.2018.5935.

7. Geisler, F.H., Coleman, W.P., Grieco, G., and Poonian, D. (2001). The Sygen multicenter acute spinal cord injury study. Spine (Phila Pa 1976) 26, S87–98.

8. Geisler, F.H., Dorsey, F.C., and Coleman, W.P. (1991). Recovery of motor function after spinal-cord injury--a randomized, placebo-controlled trial with GM-1 ganglioside. N. Engl. J. Med. 324, 1829–38.

9. Geisler, F.H., Coleman, W.P., Grieco, G., and Poonian, D. (2001). Recruitment and early treatment in a multicenter study of acute spinal cord injury. Spine (Phila. Pa. 1976). 26, S58–S67.

10. Geisler, F.H., Coleman, W.P., Grieco, G., Poonian, D., Fh, G., Wp, C., Grieco, G., Poonian, D., and Group, S.S. (2001). Measurements and recovery patterns in a multicenter study of acute spinal cord injury. Spine (Phila. Pa. 1976). 26, S68–86.

11. Kirshblum, S.C., Waring, W., Biering-Sorensen, F., Burns, S.P., Johansen, M., Schmidt-Read, M., Donovan, W., Graves, D.E., Jha, A., Jones, L., Mulcahey, M.J., and Krassioukov, A. (2011). Reference for the 2011 revision of the international standards for neurological classification of spinal cord injury. J. Spinal Cord Med..

12. Tong, B., Jutzeler, C.R., Cragg, J.J., Grassner, L., Schwab, J.M., Casha, S., Geisler, F., and Kramer, J.L.K. (2017). Serum Albumin Predicts Long-Term Neurological Outcomes After Acute Spinal Cord Injury. Neurorehabil. Neural Repair, 154596831774678.

13. Vo, A.K., Geisler, F., Grassner, L., Schwab, J., Whiteneck, G., Jutzeler, C., and Kramer, J.L.K. (2020). Serum albumin as a predictor of neurological recovery after spinal cord injury: a replication study. Spinal Cord.

14. Avila-Martin, G., Galan-Arriero, I., Gómez-Soriano, J., and Taylor, J. (2011). Treatment of rat spinal cord injury with the neurotrophic factor Albumin-Oleic acid: Translational application for paralysis, spasticity and pain. PLoS One.

15. Casha, S., Rice, T., Stirling, D.P., Silva, C., Gnanapavan, S., Giovannoni, G., John Hurlbert, R., and Wee Yong, V. (2018). Cerebrospinal Fluid Biomarkers in Human Spinal Cord Injury from a Phase II Minocycline Trial. J. Neurotrauma.

16. Ajroud-Driss, S., Saeed, M., Khan, H., Siddique, N., Hung, W.Y., Sufit, R., Heller, S., Armstrong, J., Casey, P., Siddique, T., and Lukas, T.J. (2007). Riluzole metabolism and CYP1A1/2 polymorphisms in patients with ALS. Amyotroph. Lateral Scler..

17. Bruno, R., Vivier, N., Montay, G., Le Liboux, A., Powe, L.K., Delumeau, J.C., and Rhodes, G.R. (1997). Population pharmacokinetics of riluzole in patients with amyotrophic lateral sclerosis. Clin. Pharmacol. Ther..

18. Nelis, H.J.C.F., and De Leenheer, A.P. (1982). Metabolism of minocycline in humans. Drug Metab. Dispos.

19. Saivin, S., and Houin, G. (1988). Clinical Pharmacokinetics of Doxycycline and Minocycline. Clin. Pharmacokinet..

20. Clinical, P. (2009). Guidance for Industry Drug-Induced Liver Injury?: Premarketing Clinical Evaluation. Drug Saf..

21. Watkins, P.B., Merz, M., Avigan, M.I., Kaplowitz, N., Regev, A., and Senior, J.R. (2014). The Clinical Liver Safety Assessment Best Practices Workshop: Rationale, Goals, Accomplishments and the Future. Drug Saf..

22. Ai, J.H., Xiao, F., and Hu, J.H. (2006). Guidance for industry premarketing risk assessment (I). Pharm. Care Res..

23. Hulme, C.H., Brown, S.J., Fuller, H.R., Riddell, J., Osman, A., Chowdhury, J., Kumar, N., Johnson, W.E., and Wright, K.T. (2016). The developing landscape of diagnostic and prognostic biomarkers for spinal cord injury in cerebrospinal fluid and blood. Spinal Cord, 1–12.

24. Burns, A.S., Lee, B.S., Ditunno, J.F., and Tessler, A. (2003). Patient selection for clinical trials: the reliability of the early spinal cord injury examination. J. Neurotrauma 20, 477–82.

25. Bracken, M.B., Collins, W.F., Freeman, D.F., Shepard, M.J., Wagner, F.W., Silten, R.M., Hellenbrand, K.G., Ransohoff, J., Hunt, W.E., Perot, P.L., Grossman, R.G., Green, B.A., Eisenberg, H.M., Rifkinson, N., Goodman, J.H., Meagher, J.N., Fischer, B., Clifton, G.L., Flamm, E.S., and Rawe, S.E. (1984). Efficacy of Methylprednisolone in Acute Spinal Cord Injury. JAMA J. Am. Med. Assoc..

26. Bracken, M.B., Shepard, M.J., Holford, T.R., Leo-Summers, L., Aldrich, E.F., Fazl, M., Fehlings, M., Herr, D.L., Hitchon, P.W., Marshall, L.F., Nockels, R.P., Pascale, V., Perot, P.L., Piepmeier, J., Sonntag, V.K.H., Wagner, F., Wilberger, J.E., Winn, H.R., and Young, W. (1997). Administration of methylprednisolone for 24 or 48 hours or tirilazad mesylate for 48 hours in the treatment of acute spinal cord injury: Results of the Third National Acute Spinal Cord Injury randomized controlled trial. J. Am. Med. Assoc..

27. Gutkowski, K., Chwist, A., and Hartleb, M. (2011). Liver injury induced by high-dose methylprednisolone therapy: A case report and brief review of the literature. Hepat. Mon..

28. Kadle, M.A.H., and Mazurchik, N. V. (2016). Hepatotoxicity Induced by High Dose of Methylprednisolone Therapy in a Patient with Multiple Sclerosis: A Case Report and Brief Review of Literature. Open J. Gastroenterol. 06, 146–150.

29. Shoenfeld, Y., Gurewich, Y., Gallant, L.A., and Pinkhas, J. (1981). Prednisone-induced leukocytosis. Influence of dosage, method and duration of administration on the degree of leukocytosis. Am. J. Med..

30. Vásárhelyi, B., and Debreczeni, L.A. (2017). Lab Test Findings in the Elderly. EJIFCC.

31. Fraser, C.G. (1993). Age-Related Changes in Laboratory Test Results: Clinical Implications. Drugs Aging 3, 246–257.

32. Glennie, R.A., Noonan, V.K., Fallah, N., Park, S.E., Thorogood, N.P., Cheung, A., Fisher, C.G., Dvorak, M.F., and Street, J.T. (2014). Reliability of the spine adverse events severity system (SAVES) for individuals with traumatic spinal cord injury. Spinal Cord.

33. Marion, T.E., Rivers, C.S., Kurban, D., Cheng, C.L., Fallah, N., Batke, J., Dvorak, M.F., Fisher, C.G., Kwon, B.K., Noonan, V.K., and Street, J.T. (2017). Previously Identified Common Post-Injury Adverse Events in Traumatic Spinal Cord Injury - Validation of Existing Literature and Relation to Selected Potentially Modifiable Comorbidities: A Prospective Canadian Cohort Study. J. Neurotrauma.

