## Supplementary material for "Natural Progression of Routine Laboratory Markers following Spinal Trauma: A Longitudinal, Multi-Cohort Study": Table e-1

| **Supplementary Table 1:** Subject and injury characteristics of patients *excluded* from our analysis and enrolled in the Sygen trial and Murnau study, respectively. | | | |
| --- | --- | --- | --- |
|  | **Sygen trial** | **Murnau study** | **p- value** |
| *Subject characteristics* | | |  |
| Total, n | 94 | 124 |  |
| Sex, n (%) | | | 1 |
| NA, n | 0 (0) | 0 (0) |  |
| Male | 83 (88.3) | 82 (66.1) |  |
| Female | 11 (11.7) | 42 (33.9) |  |
| Age in years at injury | | | **< 0.001** |
| NA, n | 0 | 43 |  |
| Mean ± SD | 30 ± 12 | 56 ± 22 |  |
| *Neurological/functional outcomes* | | |  |
| Baseline ASIA impairment scale, n (%) | | | **< 0.001** |
| NA | 94 (100) | 72 (58.1) |  |
| A/B/C/D | 0 (0) | 21 (16.9) |  |
| E | 0 (0) | 5 (4.0) |  |
| ND | 0 (0) | 26 (21.0) |  |
| Lower extremity motor score, mean ± SD | | |  |
| Baseline | 3.9 ± 11.4 | 32.1 ± 19.9 | **< 0.001** |
| NA, n | 0 | 68 |  |
| After one year | 13.0 ± 18.6 | 31.9 ± 21.4 | **< 0.001** |
| NA, n | 52 | 96 |  |
| *Blood markers*, n | 47 | 39 |  |
