## Supplementary material for "Natural Progression of Routine Laboratory Markers following Spinal Trauma: A Longitudinal, Multi-Cohort Study": Table e-2

| Supplementary Table 2. Hematological markers collected in the Sygen trial and Murnau Study. A total of 32 and 28 blood markers were available in the Sygen trial and Murnau study, respectively. Overall, 20 hematological markers were collected in both studies (highlighted in bold). | | |
| --- | --- | --- |
|  | **Sygen trial** | **Murnau study** |
| *Complete blood count* | | |
|  | **Erythrocytes** | **Erythrocytes** |
|  | **Hemoglobin** | **Hemoglobin** |
|  | **Hematocrit** | **Hematocrit** |
|  | **MCHC** | **MCHC** |
|  | **MCV** | **MCV** |
|  | **Thrombocytes** | **Thrombocytes** |
|  | **Leucocytes** | **Leucocytes** |
|  | Lymphocytes | Hemoglobin per erythrocyte |
|  | Monocytes |  |
|  | Neutrophils |  |
|  | Eosinophils |  |
|  | Basophils |  |
|  | MCH |  |
|  | Total serum |  |
| *Liver* | | |
|  | **Alkaline phosphatase** | **Alkaline phosphatase** |
|  | **ASAT** | **ASAT** |
|  | **ALAT** | **ALAT** |
|  | **Total bilirubin** | **Total bilirubin** |
|  | Chloride | Gamma-GT |
|  |  | Lactate dehydrogenase |
| *Kidney* | | |
|  | **Calcium** | **Calcium** |
|  | **Creatinine** | **Creatinine** |
|  | **Albumin** | **Total proteins** |
|  | **Blood urea nitrogen** | **Blood urea nitrogen** |
|  | Uric acid |  |
| *Muscle* | | |
|  | **Potassium** | **Potassium** |
|  | **Sodium** | **Sodium** |
|  |  | Cholinesterase |
| *Pancreas* | | |
|  | **Amylase** | **Amylase** |
|  |  | Lipase |
| *Others* | | |
|  | **Glucose** | **Glucose** |
|  | **Prothrombin time** | **INR** |
|  | Cholesterol | Partial thromboplasmin time |
|  | Triglycerides | CRP |
|  | Carbon dioxide | Quick test |
| *Blood markers*, n | 32 | 28 |

MCHC: mean corpuscular hemoglobin concentration; MCV: mean corpuscular volume; MCH: mean corpuscular hemoglobin; ASAT: aspartate aminotransferase; ALAT: alanine aminotransferase; Gamma-GT: gamma-glutamyl transferase; INR: international normalized ratio; CRP: protein C reactive
