## Supplementary material for "Natural Progression of Routine Laboratory Markers following Spinal Trauma: A Longitudinal, Multi-Cohort Study": Table e-3

| **Supplementary Table 3: Result of ANOVA on the hematological data from the Sygen trial.** | | | | |
| --- | --- | --- | --- | --- |
| **Hematological marker** | **df** | **Chisq** | **p- value** | **Adjusted p- value** |
| Albumin | | | | |
| Time since injury | 1 | 887 | 6.73E-195 | **4.04E-194** |
| AIS score* | 3 | 106.6 | 5.96E-23 | **3.58E-22** |
| Age | 1 | 97.59 | 5.14E-23 | **3.08E-22** |
| Neurological level | 1 | 1.008 | 3.15E-01 | 1 |
| Sex | 1 | 0.7055 | 4.01E-01 | 1 |
| Time since injury:AIS score* | 3 | 5.83 | 1.20E-01 | 0.720 |
| Alkaline phosphatase | | | | |
| Time since injury | 1 | 2.832 | 9.24E-02 | 0.554 |
| AIS score* | 3 | 28.61 | 2.70E-06 | **1.62E-05** |
| Age | 1 | 4.87 | 2.73E-02 | 0.164 |
| Neurological level | 1 | 8.881 | 2.88E-03 | **1.73E-02** |
| Sex | 1 | 0.4774 | 4.90E-01 | 1 |
| Time since injury:AIS score* | 3 | 5.257 | 1.54E-01 | 0.924 |
| Amylase | | | | |
| Time since injury | 1 | 34.99 | 3.31E-09 | **1.99E-08** |
| AIS score* | 3 | 15.33 | 1.56E-03 | **9.36E-03** |
| Age | 1 | 0.02127 | 8.84E-01 | 1 |
| Neurological level | 1 | 3.325 | 6.83E-02 | 0.410 |
| Sex | 1 | 4.744 | 2.94E-02 | 0.176 |
| Time since injury:AIS score* | 3 | 11.22 | 1.06E-02 | 0.064 |
| Prothrombin time | | | | |
| Time since injury | 1 | 194.9 | 2.78E-44 | **1.67E-43** |
| AIS score* | 3 | 3.107 | 3.75E-01 | 1 |
| Age | 1 | 0.4964 | 4.81E-01 | 1 |
| Neurological level | 1 | 0.4047 | 5.25E-01 | 1 |
| Sex | 1 | 10.89 | 9.64E-04 | **5.78E-03** |
| Time since injury:AIS score* | 3 | 0.6994 | 8.73E-01 | 1 |
| Creatinine | | | | |
| Time since injury | 1 | 8.637 | 3.29E-03 | **1.97E-02** |
| AIS score* | 3 | 10.4 | 1.55E-02 | 0.093 |
| Age | 1 | 6.727 | 9.49E-03 | 0.057 |
| Neurological level | 1 | 1.583 | 2.08E-01 | 1 |
| Sex | 1 | 79.83 | 4.08E-19 | **2.45E-18** |
| Time since injury:AIS score* | 3 | 6.6 | 8.58E-02 | 0.515 |
| Bilirubin | | | | |
| Time since injury | 1 | 19.02 | 1.29E-05 | **7.74E-05** |
| AIS score* | 3 | 7.131 | 6.78E-02 | 0.407 |
| Age | 1 | 2.31 | 1.29E-01 | 0.774 |
| Neurological level | 1 | 0.6454 | 4.22E-01 | 1 |
| Sex | 1 | 3.405 | 6.50E-02 | 0.390 |
| Time since injury:AIS score* | 3 | 3.2 | 3.62E-01 | 1 |
| Uric acid | | | | |
| Time since injury | 1 | 422.5 | 7.00E-94 | **4.20E-93** |
| AIS score* | 3 | 12.06 | 7.19E-03 | **4.31E-02** |
| Age | 1 | 4.838 | 2.78E-02 | 0.167 |
| Neurological level | 1 | 0.04564 | 8.31E-01 | 1 |
| Sex | 1 | 78.52 | 7.92E-19 | **4.75E-18** |
| Time since injury:AIS score* | 3 | 25.28 | 1.35E-05 | **8.10E-05** |
| Blood urea nitrogen | | | | |
| Time since injury | 1 | 93.12 | 4.92E-22 | **2.95E-21** |
| AIS score* | 3 | 7.98 | 4.64E-02 | 0.278 |
| Age | 1 | 48.02 | 4.22E-12 | **2.53E-11** |
| Neurological level | 1 | 0.7502 | 3.86E-01 | 1 |
| Sex | 1 | 59.6 | 1.16E-14 | **6.96E-14** |
| Time since injury:AIS score* | 3 | 2.812 | 4.21E-01 | 1 |
| Calcium | | | | |
| Time since injury | 1 | 493.6 | 2.31E-109 | **1.39E-108** |
| AIS score* | 3 | 40.84 | 7.06E-09 | **4.24E-08** |
| Age | 1 | 70.41 | 4.82E-17 | **2.89E-16** |
| Neurological level | 1 | 4.869 | 2.73E-02 | 0.164 |
| Sex | 1 | 0.217 | 6.41E-01 | 1 |
| Time since injury:AIS score* | 3 | 6.786 | 7.91E-02 | 0.475 |
| MCHC | | | | |
| Time since injury | 1 | 14.22 | 1.63E-04 | **9.78E-04** |
| AIS score* | 3 | 1.645 | 6.49E-01 | 1 |
| Age | 1 | 4.984 | 2.56E-02 | 0.154 |
| Neurological level | 1 | 0.0616 | 8.04E-01 | 1 |
| Sex | 1 | 1.043 | 3.07E-01 | 1 |
| Time since injury:AIS score* | 3 | 8.037 | 4.53E-02 | 0.272 |
| Cholesterol | | | | |
| Time since injury | 1 | 210.2 | 1.26E-47 | **7.56E-47** |
| AIS score* | 3 | 31.62 | 6.29E-07 | **3.77E-06** |
| Age | 1 | 33.69 | 6.45E-09 | **3.87E-08** |
| Neurological level | 1 | 6.217 | 1.27E-02 | 0.076 |
| Sex | 1 | 12 | 5.31E-04 | **3.19E-03** |
| Time since injury:AIS score* | 3 | 10.04 | 1.82E-02 | 0.109 |
| Creatin phosphokinase | | | | |
| Time since injury | 1 | 76.41 | 2.31E-18 | **1.39E-17** |
| AIS score* | 3 | 7.315 | 6.25E-02 | 0.375 |
| Age | 1 | 15.35 | 8.94E-05 | **5.36E-04** |
| Neurological level | 1 | 1.158 | 2.82E-01 | 1 |
| Sex | 1 | 15.77 | 7.15E-05 | **4.29E-04** |
| Time since injury:AIS score* | 3 | 2.616 | 4.55E-01 | 1 |
| Chloride | | | | |
| Time since injury | 1 | 18.66 | 1.56E-05 | **9.36E-05** |
| AIS score* | 3 | 1.264 | 7.38E-01 | 1 |
| Age | 1 | 0.1989 | 6.56E-01 | 1 |
| Neurological level | 1 | 4.02 | 4.50E-02 | 0.270 |
| Sex | 1 | 14.2 | 1.65E-04 | **9.90E-04** |
| Time since injury:AIS score* | 3 | 0.6082 | 8.95E-01 | 1 |
| Carbon dioxide | | | | |
| Time since injury | 1 | 10.32 | 1.32E-03 | **7.92E-03** |
| AIS score* | 3 | 17.72 | 5.03E-04 | **3.02E-03** |
| Age | 1 | 0.4867 | 4.85E-01 | 1 |
| Neurological level | 1 | 1.143 | 2.85E-01 | 1 |
| Sex | 1 | 4.605 | 3.19E-02 | 0.191 |
| Time since injury:AIS score* | 3 | 2.486 | 4.78E-01 | 1 |
| Neutrophils | | | | |
| Time since injury | 1 | 458.1 | 1.22E-101 | **7.32E-101** |
| AIS score* | 3 | 10.63 | 1.39E-02 | 0.083 |
| Age | 1 | 12.26 | 4.62E-04 | **2.77E-03** |
| Neurological level | 1 | 0.0002821 | 9.87E-01 | 1 |
| Sex | 1 | 2.65 | 1.04E-01 | 0.624 |
| Time since injury:AIS score* | 3 | 2.321 | 5.08E-01 | 1 |
| Lymphocytes | | | | |
| Time since injury | 1 | 547.4 | 4.63E-121 | **2.78E-120** |
| AIS score* | 3 | 19.59 | 2.06E-04 | **1.24E-03** |
| Age | 1 | 18.09 | 2.10E-05 | **1.26E-04** |
| Neurological level | 1 | 0.3279 | 5.67E-01 | 1 |
| Sex | 1 | 8.003 | 4.67E-03 | **2.80E-02** |
| Time since injury:AIS score* | 3 | 1.117 | 7.73E-01 | 1 |
| Monocytes | | | | |
| Time since injury | 1 | 49.95 | 1.58E-12 | **9.48E-12** |
| AIS score* | 3 | 0.4049 | 9.39E-01 | 1 |
| Age | 1 | 2.999 | 8.33E-02 | 0.500 |
| Neurological level | 1 | 0.4214 | 5.16E-01 | 1 |
| Sex | 1 | 2.108 | 1.47E-01 | 0.882 |
| Time since injury:AIS score* | 3 | 5.303 | 1.51E-01 | 0.906 |
| Eosinophils | | | | |
| Time since injury | 1 | 117.1 | 2.75E-27 | **1.65E-26** |
| AIS score* | 3 | 3.06 | 3.82E-01 | 1 |
| Age | 1 | 1.66 | 1.98E-01 | 1 |
| Neurological level | 1 | 1.276 | 2.59E-01 | 1 |
| Sex | 1 | 0.2558 | 6.13E-01 | 1 |
| Time since injury:AIS score* | 3 | 7.908 | 4.79E-02 | 0.287 |
| Basophils | | | | |
| Time since injury | 1 | 28.1 | 1.15E-07 | **6.90E-07** |
| AIS score* | 3 | 5.442 | 1.42E-01 | 0.52 |
| Age | 1 | 6.167 | 1.30E-02 | 0.078 |
| Neurological level | 1 | 0.1958 | 6.58E-01 | 1 |
| Sex | 1 | 2.029 | 1.54E-01 | 0.924 |
| Time since injury:AIS score* | 3 | 1.946 | 5.84E-01 | 1 |
| Glucose | | | | |
| Time since injury | 1 | 150.1 | 1.62E-34 | **9.72E-34** |
| AIS score* | 3 | 8.943 | 3.01E-02 | 0.181 |
| Age | 1 | 96.28 | 1.00E-22 | **6.00E-22** |
| Neurological level | 1 | 0.1352 | 7.13E-01 | 1 |
| Sex | 1 | 1.02 | 3.13E-01 | 1 |
| Time since injury:AIS score* | 3 | 0.2356 | 9.72E-01 | 1 |
| Hematocrit | | | | |
| Time since injury | 1 | 900.4 | 7.84E-198 | **4.70E-197** |
| AIS score* | 3 | 77.1 | 1.29E-16 | **7.74E-16** |
| Age | 1 | 24.44 | 7.67E-07 | **4.60E-06** |
| Neurological level | 1 | 1.222 | 2.69E-01 | 1 |
| Sex | 1 | 37.65 | 8.45E-10 | **5.07E-09** |
| Time since injury:AIS score* | 3 | 9.875 | 1.97E-02 | 0.118 |
| Hemoglobin | | | | |
| Time since injury | 1 | 907.7 | 2.04E-199 | **1.22E-198** |
| AIS score* | 3 | 77.89 | 8.72E-17 | **5.23E-16** |
| Age | 1 | 27.88 | 1.29E-07 | **7.74E-07** |
| Neurological level | 1 | 1.142 | 2.85E-01 | 1 |
| Sex | 1 | 39 | 4.24E-10 | **2.54E-09** |
| Time since injury:AIS score* | 3 | 8.592 | 3.52E-02 | 0.211 |
| Potassium | | | | |
| Time since injury | 1 | 1.379 | 2.40E-01 | 1 |
| AIS score* | 3 | 3.692 | 2.97E-01 | 1 |
| Age | 1 | 0.04286 | 8.36E-01 | 1 |
| Neurological level | 1 | 0.6681 | 4.14E-01 | 1 |
| Sex | 1 | 2.35 | 1.25E-01 | 0.750 |
| Time since injury:AIS score* | 3 | 1.661 | 6.46E-01 | 1 |
| MCH | | | | |
| Time since injury | 1 | 140.3 | 2.34E-32 | **1.40E-31** |
| AIS score* | 3 | 5.171 | 1.60E-01 | 0.960 |
| Age | 1 | 9.149 | 2.49E-03 | **1.49E-02** |
| Neurological level | 1 | 0.001352 | 9.71E-01 | 1 |
| Sex | 1 | 0.4363 | 5.09E-01 | 1 |
| Time since injury:AIS score* | 3 | 12.53 | 5.77E-03 | **3.46E-02** |
| MCV | | | | |
| Time since injury | 1 | 1.827 | 1.77E-01 | 1 |
| AIS score* | 3 | 4.953 | 1.75E-01 | 1 |
| Age | 1 | 2.174 | 1.40E-01 | 0.840 |
| Neurological level | 1 | 1.41 | 2.35E-01 | 1 |
| Sex | 1 | 2.632 | 1.05E-01 | 0.630 |
| Time since injury:AIS score* | 3 | 2.813 | 4.21E-01 | 1 |
| Sodium | | | | |
| Time since injury | 1 | 54.37 | 1.66E-13 | **9.96E-13** |
| AIS score* | 3 | 20.5 | 1.34E-04 | **8.04E-04** |
| Age | 1 | 9.316 | 2.27E-03 | **1.36E-02** |
| Neurological level | 1 | 2.436 | 1.19E-01 | 0.714 |
| Sex | 1 | 0.1578 | 6.91E-01 | 1 |
| Time since injury:AIS score* | 3 | 0.7529 | 8.61E-01 | 1 |
| Thrombocytes | | | | |
| Time since injury | 1 | 0.1253 | 7.23E-01 | 1 |
| AIS score* | 3 | 5.794 | 1.22E-01 | 0.732 |
| Age | 1 | 1.804 | 1.79E-01 | 1 |
| Neurological level | 1 | 3.551 | 5.95E-02 | 0.357 |
| Sex | 1 | 1.062 | 3.03E-01 | 1 |
| Time since injury:AIS score* | 3 | 1.999 | 5.73E-01 | 1 |
| Erythrocytes | | | | |
| Time since injury | 1 | 1026 | 4.56E-225 | **2.74E-224** |
| AIS score* | 3 | 62.36 | 1.84E-13 | **1.10E-12** |
| Age | 1 | 46.56 | 8.89E-12 | **5.33E-11** |
| Neurological level | 1 | 1.184 | 2.77E-01 | 1 |
| Sex | 1 | 34.02 | 5.45E-09 | **3.27E-08** |
| Time since injury:AIS score* | 3 | 12.23 | 6.64E-03 | **3.98E-02** |
| ASAT | | | | |
| Time since injury | 1 | 104.2 | 1.80E-24 | **1.08E-23** |
| AIS score* | 3 | 15.75 | 1.28E-03 | **7.68E-03** |
| Age | 1 | 1.31 | 2.52E-01 | 1 |
| Neurological level | 1 | 0.213 | 6.44E-01 | 1 |
| Sex | 1 | 17.49 | 2.89E-05 | **1.73E-04** |
| Time since injury:AIS score* | 3 | 6.706 | 8.19E-02 | 0.491 |
| ALAT | | | | |
| Time since injury | 1 | 110.6 | 7.06E-26 | **4.24E-25** |
| AIS score* | 3 | 12.81 | 5.06E-03 | **3.04E-02** |
| Age | 1 | 0.8431 | 3.59E-01 | 1 |
| Neurological level | 1 | 0.07282 | 7.87E-01 | 1 |
| Sex | 1 | 15.15 | 9.94E-05 | **5.96E-04** |
| Time since injury:AIS score* | 3 | 3.958 | 2.66E-01 | 1 |
| Total serum | | | | |
| Time since injury | 1 | 1022 | 3.60E-224 | **2.16E-223** |
| AIS score* | 3 | 47.61 | 2.58E-10 | **1.55E-09** |
| Age | 1 | 45.5 | 1.53E-11 | **9.18E-11** |
| Neurological level | 1 | 3.658 | 5.58E-02 | 0.335 |
| Sex | 1 | 4.1 | 4.29E-02 | 0.257 |
| Time since injury:AIS score* | 3 | 8.327 | 3.97E-02 | 0.238 |
| Triglycerides | | | | |
| Time since injury | 1 | 29.6 | 5.32E-08 | **3.19E-07** |
| AIS score* | 3 | 5.013 | 1.71E-01 | 1 |
| Age | 1 | 52.33 | 4.69E-13 | **2.81E-12** |
| Neurological level | 1 | 1.962 | 1.61E-01 | 0.966 |
| Sex | 1 | 0.9648 | 3.26E-01 | 1 |
| Time since injury:AIS score* | 3 | 9.396 | 2.45E-02 | 0.147 |
| Leucocytes | | | | |
| Time since injury | 1 | 251 | 1.54E-56 | **9.24E-56** |
| AIS score* | 3 | 6.89 | 7.55E-02 | 0.453 |
| Age | 1 | 4.124 | 4.23E-02 | 0.254 |
| Neurological level | 1 | 0.4389 | 5.08E-01 | 1 |
| Sex | 1 | 2.137 | 1.44E-01 | 0.864 |
| Time since injury:AIS score* | 3 | 2.18 | 5.36E-01 | 1 |

*ASIA impairment scale: A, no sensory or motor function is preserved; B, sensory function is preserved below the level of the injury, but there is no motor function; C, motor function is preserved below the neurological level, and more than half of the key muscles below the neurological level have a muscle grade of <3; D, motor function is preserved below the neurological level, and at least half of the key muscles below the neurological level have a muscle grade of >3.

Neurological level of injury is a binary variable defined as injury above/at T6 or below T6.

Abbreviations: MCHC: mean corpuscular hemoglobin concentration; MCH: mean corpuscular hemoglobin; MCV: mean corpuscular volume; ASAT: aspartate aminotransferase; ALAT: alanine aminotransferase
