## Supplementary material for "Natural Progression of Routine Laboratory Markers following Spinal Trauma: A Longitudinal, Multi-Cohort Study": Table e-4

| **Supplementary Table 4: Result of ANOVA on the hematological data from the Murnau study.** | | | | |
| --- | --- | --- | --- | --- |
| **Hematological marker** | **df** | **Chisq** | **p- value** | **Adjusted p- value** |
| Amylase | | | | |
| Time since injury | 1 | 3.169 | 7.50E-02 | 0.450 |
| AIS score* | 3 | 13.77 | 3.23E-03 | **1.94E-02** |
| Age | 1 | 1.984 | 1.59E-01 | 0.954 |
| Neurological level | 1 | 0.04816 | 8.26E-01 | 1 |
| Sex | 1 | 1.898 | 1.68E-01 | 1 |
| Time since injury:AIS score* | 3 | 2.326 | 5.08E-01 | 1 |
| Alkaline phosphatase | | | | |
| Time since injury | 1 | 74.62 | 5.70E-18 | **3.42E-17** |
| AIS score* | 3 | 0.3214 | 9.56E-01 | 1 |
| Age | 1 | 7.797 | 5.23E-03 | **3.14E-02** |
| Neurological level | 1 | 1.102 | 2.94E-01 | 1 |
| Sex | 1 | 0.0005382 | 9.81E-01 | 1 |
| Time since injury:AIS score* | 3 | 9.533 | 2.30E-02 | 0.138 |
| Calcium | | | | |
| Time since injury | 1 | 215.8 | 7.32E-49 | **4.39E-48** |
| AIS score* | 3 | 23.84 | 2.70E-05 | **1.62E-04** |
| Age | 1 | 7.246 | 7.11E-03 | **4.27E-02** |
| Neurological level | 1 | 3.694 | 5.46E-02 | 0.328 |
| Sex | 1 | 0.0002722 | 9.87E-01 | 1 |
| Time since injury:AIS score* | 3 | 18.61 | 3.29E-04 | **1.97E-03** |
| Cholinesterase | | | | |
| Time since injury | 1 | 18.93 | 1.36E-05 | **8.16E-05** |
| AIS score* | 3 | 51.81 | 3.29E-11 | **1.97E-10** |
| Age | 1 | 9.521 | 2.03E-03 | **1.22E-02** |
| Neurological level | 1 | 0.5496 | 4.58E-01 | 1 |
| Sex | 1 | 2.284 | 1.31E-01 | 0.786 |
| Time since injury:AIS score* | 3 | 17.3 | 6.12E-04 | **3.67E-03** |
| Creatinine | | | | |
| Time since injury | 1 | 92.59 | 6.42E-22 | **3.85E-21** |
| AIS score* | 3 | 0.3007 | 9.60E-01 | 1 |
| Age | 1 | 23 | 1.62E-06 | **9.72E-06** |
| Neurological level | 1 | 0.6399 | 4.24E-01 | 1 |
| Sex | 1 | 40.44 | 2.02E-10 | **1.21E-09** |
| Time since injury:AIS score* | 3 | 36.95 | 4.72E-08 | **2.83E-07** |
| CRP | | | | |
| Time since injury | 1 | 88.82 | 4.32E-21 | **2.59E-20** |
| AIS score* | 3 | 37.45 | 3.70E-08 | **2.22E-07** |
| Age | 1 | 6.507 | 1.07E-02 | 0.064 |
| Neurological level | 1 | 0.6665 | 4.14E-01 | 1 |
| Sex | 1 | 0.01262 | 9.11E-01 | 1 |
| Time since injury:AIS score* | 3 | 12.19 | 6.74E-03 | **4.04E-02** |
| Erythrocytes | | | | |
| Time since injury | 1 | 278 | 2.05E-62 | **1.23E-61** |
| AIS score* | 3 | 98.34 | 3.54E-21 | **2.12E-20** |
| Age | 1 | 19.46 | 1.03E-05 | **6.18E-05** |
| Neurological level | 1 | 0.07407 | 7.85E-01 | 1 |
| Sex | 1 | 6.324 | 1.19E-02 | 0.071 |
| Time since injury:AIS score* | 3 | 48.74 | 1.48E-10 | **8.88E-10** |
| Gamma GT | | | | |
| Time since injury | 1 | 1.101 | 2.94E-01 | 1 |
| AIS score* | 3 | 6.364 | 9.52E-02 | 0.571 |
| Age | 1 | 4.966 | 2.59E-02 | 0.155 |
| Neurological level | 1 | 8.502 | 3.55E-03 | **2.13E-02** |
| Sex | 1 | 0.9434 | 3.31E-01 | 1 |
| Time since injury:AIS score* | 3 | 10.55 | 1.45E-02 | 0.087 |
| Bilirubin | | | | |
| Time since injury | 1 | 17.04 | 3.66E-05 | **2.20E-04** |
| AIS score* | 3 | 2.099 | 5.52E-01 | 1 |
| Age | 1 | 1.452 | 2.28E-01 | 1 |
| Neurological level | 1 | 3.501 | 6.13E-02 | 0.368 |
| Sex | 1 | 0.932 | 3.34E-01 | 1 |
| Time since injury:AIS score* | 3 | 7.554 | 5.62E-02 | 0.337 |
| Total proteins | | | | |
| Time since injury | 1 | 224 | 1.24E-50 | **7.44E-50** |
| AIS score* | 3 | 49.19 | 1.19E-10 | **7.14E-10** |
| Age | 1 | 21.29 | 3.95E-06 | **2.37E-05** |
| Neurological level | 1 | 0.1201 | 7.29E-01 | 1 |
| Sex | 1 | 2.905 | 8.83E-02 | 0.530 |
| Time since injury:AIS score* | 3 | 21.53 | 8.16E-05 | **4.90E-04** |
| Glucose | | | | |
| Time since injury | 1 | 6.26 | 1.24E-02 | 0.074 |
| AIS score* | 3 | 4.322 | 2.29E-01 | 1 |
| Age | 1 | 2.692 | 1.01E-01 | 0.606 |
| Neurological level | 1 | 2.463 | 1.17E-01 | 0.702 |
| Sex | 1 | 0.02687 | 8.70E-01 | 1 |
| Time since injury:AIS score* | 3 | 2.013 | 5.70E-01 | 1 |
| Blood urea | | | | |
| Time since injury | 1 | 126.9 | 1.98E-29 | **1.19E-28** |
| AIS score* | 3 | 8.439 | 3.78E-02 | 0.227 |
| Age | 1 | 57.87 | 2.80E-14 | **1.68E-13** |
| Neurological level | 1 | 5.241 | 2.21E-02 | 0.133 |
| Sex | 1 | 21.18 | 4.19E-06 | **2.51E-05** |
| Time since injury:AIS score* | 3 | 25.9 | 1.00E-05 | **6.00E-05** |
| Hemoglobin | | | | |
| Time since injury | 1 | 147.7 | 5.57E-34 | **3.34E-33** |
| AIS score* | 3 | 91.79 | 9.03E-20 | **5.42E-19** |
| Age | 1 | 9.713 | 1.83E-03 | **1.10E-02** |
| Neurological level | 1 | 0.03842 | 8.45E-01 | 1 |
| Sex | 1 | 8.339 | 3.88E-03 | **2.33E-02** |
| Time since injury:AIS score* | 3 | 27.55 | 4.52E-06 | **2.71E-05** |
| Hemoglobin per erythrocyte | | | | |
| Time since injury | 1 | 357.8 | 8.46E-80 | **5.08E-79** |
| AIS score* | 3 | 1.12 | 7.72E-01 | 1 |
| Age | 1 | 11.61 | 6.54E-04 | **3.92E-03** |
| Neurological level | 1 | 0.4882 | 4.85E-01 | 1 |
| Sex | 1 | 0.4735 | 4.91E-01 | 1 |
| Time since injury:AIS score* | 3 | 54.96 | 7.01E-12 | **4.21E-11** |
| Hematocrit | | | | |
| Time since injury | 1 | 217.4 | 3.31E-49 | **1.99E-48** |
| AIS score* | 3 | 96.27 | 9.86E-21 | **5.92E-20** |
| Age | 1 | 11.2 | 8.17E-04 | **4.90E-03** |
| Neurological level | 1 | 0.1286 | 7.20E-01 | 1 |
| Sex | 1 | 5.939 | 1.48E-02 | 0.089 |
| Time since injury:AIS score* | 3 | 36.56 | 5.69E-08 | **3.41E-07** |
| INR | | | | |
| Time since injury | 1 | 12.65 | 3.76E-04 | **2.26E-03** |
| AIS score* | 3 | 2.655 | 4.48E-01 | 1 |
| Age | 1 | 2.502 | 1.14E-01 | 0.684 |
| Neurological level | 1 | 0.3038 | 5.82E-01 | 1 |
| Sex | 1 | 0.06602 | 7.97E-01 | 1 |
| Time since injury:AIS score* | 3 | 26.36 | 8.00E-06 | **4.80E-05** |
| Potassium | | | | |
| Time since injury | 1 | 31.27 | 2.24E-08 | **1.34E-07** |
| AIS score* | 3 | 12.25 | 6.56E-03 | **3.94E-02** |
| Age | 1 | 1.024 | 3.12E-01 | 1 |
| Neurological level | 1 | 0.08262 | 7.74E-01 | 1 |
| Sex | 1 | 12.89 | 3.30E-04 | **1.98E-03** |
| Time since injury:AIS score* | 3 | 18.24 | 3.92E-04 | **2.35E-03** |
| Lactate dehydrogenase | | | | |
| Time since injury | 1 | 128.1 | 1.09E-29 | **6.54E-29** |
| AIS score* | 3 | 9.469 | 2.37E-02 | 0.142 |
| Age | 1 | 1.66 | 1.98E-01 | 1 |
| Neurological level | 1 | 17.56 | 2.79E-05 | **1.67E-04** |
| Sex | 1 | 0.2117 | 6.45E-01 | 1 |
| Time since injury:AIS score* | 3 | 4.976 | 1.74E-01 | 1 |
| Leucocytes | | | | |
| Time since injury | 1 | 67.62 | 1.99E-16 | **1.19E-15** |
| AIS score* | 3 | 3.091 | 3.78E-01 | 1 |
| Age | 1 | 0.1409 | 7.07E-01 | 1 |
| Neurological level | 1 | 3.188 | 7.42E-02 | 0.445 |
| Sex | 1 | 0.005449 | 9.41E-01 | 1 |
| Time since injury:AIS score* | 3 | 3.22 | 3.59E-01 | 1 |
| Lipase | | | | |
| Time since injury | 1 | 0.002547 | 9.60E-01 | 1 |
| AIS score* | 3 | 7.548 | 5.63E-02 | 0.338 |
| Age | 1 | 5.734 | 1.66E-02 | 0.100 |
| Neurological level | 1 | 5.688 | 1.71E-02 | 0.103 |
| Sex | 1 | 1.183 | 2.77E-01 | 1 |
| Time since injury:AIS score* | 3 | 3.104 | 3.76E-01 | 1 |
| MCHC | | | | |
| Time since injury | 1 | 172.7 | 1.94E-39 | **1.16E-38** |
| AIS score* | 3 | 4.72 | 1.93E-01 | 1 |
| Age | 1 | 3.84E-05 | 9.95E-01 | 1 |
| Neurological level | 1 | 0.1445 | 7.04E-01 | 1 |
| Sex | 1 | 7.649 | 5.68E-03 | **3.41E-02** |
| Time since injury:AIS score* | 3 | 17.26 | 6.24E-04 | **3.74E-03** |
| MCV | | | | |
| Time since injury | 1 | 74.72 | 5.42E-18 | **3.25E-17** |
| AIS score* | 3 | 2.303 | 5.12E-01 | 1 |
| Age | 1 | 15.14 | 9.97E-05 | **5.98E-04** |
| Neurological level | 1 | 0.000624 | 9.80E-01 | 1 |
| Sex | 1 | 0.3478 | 5.55E-01 | 1 |
| Time since injury:AIS score* | 3 | 24.31 | 2.16E-05 | **1.30E-04** |
| Sodium | | | | |
| Time since injury | 1 | 49.84 | 1.67E-12 | **1.00E-11** |
| AIS score* | 3 | 0.07835 | 9.94E-01 | 1 |
| Age | 1 | 5.029 | 2.49E-02 | 0.149 |
| Neurological level | 1 | 0.02276 | 8.80E-01 | 1 |
| Sex | 1 | 0.8982 | 3.43E-01 | 1 |
| Time since injury:AIS score* | 3 | 15.3 | 1.58E-03 | **9.48E-03** |
| Prothrombin time | | | | |
| Time since injury | 1 | 15.25 | 9.42E-05 | **5.65E-04** |
| AIS score* | 3 | 20.2 | 1.55E-04 | **9.30E-04** |
| Age | 1 | 0.5929 | 4.41E-01 | 1 |
| Neurological level | 1 | 0.7423 | 3.89E-01 | 1 |
| Sex | 1 | 6.689 | 9.70E-03 | 0.058 |
| Time since injury:AIS score* | 3 | 7.77 | 5.10E-02 | 0.306 |
| Quick test | | | | |
| Time since injury | 1 | 45.21 | 1.77E-11 | **1.06E-10** |
| AIS score* | 3 | 4.906 | 1.79E-01 | 1 |
| Age | 1 | 4.365 | 3.67E-02 | 0.220 |
| Neurological level | 1 | 1.142 | 2.85E-01 | 1 |
| Sex | 1 | 0.7402 | 3.90E-01 | 1 |
| Time since injury:AIS score* | 3 | 23.11 | 3.83E-05 | **2.30E-04** |
| Thrombocytes | | | | |
| Time since injury | 1 | 40.07 | 2.45E-10 | **1.47E-09** |
| AIS score* | 3 | 6.141 | 1.05E-01 | 0.630 |
| Age | 1 | 5.781 | 1.62E-02 | 0.097 |
| Neurological level | 1 | 5.423 | 1.99E-02 | 0.119 |
| Sex | 1 | 0.6402 | 4.24E-01 | 1 |
| Time since injury:AIS score* | 3 | 2.662 | 4.47E-01 | 1 |
| ASAT | | | | |
| Time since injury | 1 | 30.19 | 3.91E-08 | **2.35E-07** |
| AIS score* | 3 | 11.32 | 1.01E-02 | 0.061 |
| Age | 1 | 0.02795 | 8.67E-01 | 1 |
| Neurological level | 1 | 0.2564 | 6.13E-01 | 1 |
| Sex | 1 | 0.6442 | 4.22E-01 | 1 |
| Time since injury:AIS score* | 3 | 4.843 | 1.84E-01 | 1 |
| ALAT | | | | |
| Time since injury | 1 | 4.38 | 3.64E-02 | 0.218 |
| AIS score* | 3 | 7.634 | 5.42E-02 | 0.325 |
| Age | 1 | 2.632 | 1.05E-01 | 0.630 |
| Neurological level | 1 | 8.411 | 3.73E-03 | **2.24E-02** |
| Sex | 1 | 0.6091 | 4.35E-01 | 1 |
| Time since injury:AIS score* | 3 | 1.904 | 5.93E-01 | 1 |

*ASIA impairment scale: A, no sensory or motor function is preserved; B, sensory function is preserved below the level of the injury, but there is no motor function; C, motor function is preserved below the neurological level, and more than half of the key muscles below the neurological level have a muscle grade of <3; D, motor function is preserved below the neurological level, and at least half of the key muscles below the neurological level have a muscle grade of >3.

Neurological level of injury is a binary variable defined as injury above/at T6 or below T6.

**Abbreviations:** CRP: C-reactive protein; Gamma GT: gamma-glutamyl transferase; INR: international normalised ratio; MCHC: mean corpuscular hemoglobin concentration; MCV: mean corpuscular volume; ASAT: aspartate aminotransferase; ALAT: alanine aminotransferase
