## Supplementary material for "Natural Progression of Routine Laboratory Markers following Spinal Trauma: A Longitudinal, Multi-Cohort Study": Table e-5

| **Supplementary Table 5: Result of pairwise comparisons between the AIS grades on the hematological data from the Sygen study.** | | | |
| --- | --- | --- | --- |
| **Hematological marker** | **Effect** | **p- value** | **Adjusted p- value** |
| Albumin | | | |
| AIS score* (AIS) A – AIS B | 0.192 | 4.70E-06 | **1.73E-05** |
| AIS A – AIS C | 0.261 | 8.88E-16 | **5.22E-15** |
| AIS A – AIS D | 0.446 | 2.66E-12 | **8.05E-12** |
| AIS B – AIS C | 0.069 | 0.147 | 0.449 |
| AIS B – AIS D | 0.254 | 4.70E-04 | **2.16E-03** |
| AIS C – AIS D | 0.185 | 6.12E-03 | **2.83E-02** |
| Alkaline phosphatase | | | |
| AIS A – AIS B | -25.22 | 2.18E-04 | **1.18E-03** |
| AIS A – AIS C | -24.408 | 3.88E-06 | **1.76E-05** |
| AIS A – AIS D | -23.908 | 2.08E-02 | 8.76E-02 |
| AIS B – AIS C | 0.812 | 0.917 | 1 |
| AIS B – AIS D | 1.312 | 0.911 | 0.999 |
| AIS C – AIS D | 0.5 | 0.964 | 1 |
| Amylase | | | |
| AIS A – AIS B | -9.788 | 0.175 | 0.509 |
| AIS A – AIS C | -13.187 | 1.85E-02 | 7.88E-02 |
| AIS A – AIS D | -22.952 | 3.66E-02 | 0.145 |
| AIS B – AIS C | -3.398 | 0.678 | 0.974 |
| AIS B – AIS D | -13.163 | 0.292 | 0.703 |
| AIS C – AIS D | -9.765 | 0.400 | 0.824 |
| Prothrombin time | | | |
| AIS A – AIS B | 0.605 | 0.252 | 0.645 |
| AIS A – AIS C | -0.173 | 0.674 | 0.973 |
| AIS A – AIS D | -1.215 | 0.132 | 0.414 |
| AIS B – AIS C | -0.779 | 0.195 | 0.548 |
| AIS B – AIS D | -1.821 | 4.72E-02 | 0.181 |
| AIS C – AIS D | -1.042 | 0.222 | 0.596 |
| Creatinine | | | |
| AIS A – AIS B | 0.064 | 4.55E-02 | 0.175 |
| AIS A – AIS C | 0.065 | 7.99E-03 | **3.58E-02** |
| AIS A – AIS D | 0.152 | 1.68E-03 | **7.81E-03** |
| AIS B – AIS C | 0.002 | 0.961 | 1 |
| AIS B – AIS D | 0.088 | 0.110 | 0.360 |
| AIS C – AIS D | 0.086 | 0.0913 | 0.312 |
| Total bilirubin | | | |
| AIS A – AIS B | -0.061 | 0.46 | 0.873 |
| AIS A – AIS C | -0.129 | 4.40E-02 | 0.170 |
| AIS A – AIS D | -0.204 | 0.106 | 0.351 |
| AIS B – AIS C | -0.068 | 0.469 | 0.880 |
| AIS B – AIS D | -0.143 | 0.320 | 0.739 |
| AIS C – AIS D | -0.075 | 0.575 | 0.939 |
| Uric acid | | | |
| AIS A – AIS B | 0.031 | 0.830 | 0.996 |
| AIS A – AIS C | 0.146 | 0.187 | 0.532 |
| AIS A – AIS D | 0.517 | 1.77E-02 | 7.58E-02 |
| AIS B – AIS C | 0.116 | 0.477 | 0.885 |
| AIS B – AIS D | 0.486 | 5.02E-02 | 0.190 |
| AIS C – AIS D | 0.37 | 0.108 | 0.357 |
| Blood urea | | | |
| AIS A – AIS B | -0.605 | 0.421 | 0.843 |
| AIS A – AIS C | -1.033 | 7.67E-02 | 0.271 |
| AIS A – AIS D | -2.18 | 5.66E-02 | 0.211 |
| AIS B – AIS C | -0.428 | 0.617 | 0.956 |
| AIS B – AIS D | -1.575 | 0.227 | 0.604 |
| AIS C – AIS D | -1.147 | 0.343 | 0.766 |
| Calcium | | | |
| AIS A – AIS B | 0.129 | 2.17E-02 | 9.08E-02 |
| AIS A – AIS C | 0.214 | 9.08E-07 | **2.86E-06** |
| AIS A – AIS D | 0.317 | 1.99E-04 | **9.07E-04** |
| AIS B – AIS C | 0.085 | 0.183 | 0.525 |
| AIS B – AIS D | 0.188 | 5.29E-02 | 0.199 |
| AIS C – AIS D | 0.103 | 0.253 | 0.646 |
| MCHC | | | |
| AIS A – AIS B | -0.023 | 0.773 | 0.991 |
| AIS A – AIS C | 0.061 | 0.323 | 0.743 |
| AIS A – AIS D | 0.125 | 0.304 | 0.719 |
| AIS B – AIS C | 0.084 | 0.352 | 0.776 |
| AIS B – AIS D | 0.148 | 0.285 | 0.693 |
| AIS C – AIS D | 0.064 | 0.621 | 0.957 |
| Cholesterol | | | |
| AIS A – AIS B | 0.439 | 0.908 | 0.999 |
| AIS A – AIS C | 11.359 | 1.23E-04 | **5.76E-04** |
| AIS A – AIS D | 22.312 | 1.22E-04 | **6.35E-04** |
| AIS B – AIS C | 10.919 | 1.17E-02 | 5.13E-02 |
| AIS B – AIS D | 21.873 | 9.38E-04 | **4.70E-03** |
| AIS C – AIS D | 10.953 | 7.45E-02 | 0.265 |
| Creatin phosphokinase | | | |
| AIS A – AIS B | -152.119 | 8.83E-02 | 0.304 |
| AIS A – AIS C | -82.143 | 0.234 | 0.616 |
| AIS A – AIS D | -184.929 | 0.168 | 0.495 |
| AIS B – AIS C | 69.976 | 0.489 | 0.894 |
| AIS B – AIS D | -32.81 | 0.831 | 0.996 |
| AIS C – AIS D | -102.786 | 0.469 | 0.880 |
| Chloride | | | |
| AIS A – AIS B | -0.079 | 0.799 | 0.994 |
| AIS A – AIS C | 0.205 | 0.396 | 0.820 |
| AIS A – AIS D | 0.34 | 0.470 | 0.881 |
| AIS B – AIS C | 0.284 | 0.421 | 0.843 |
| AIS B – AIS D | 0.42 | 0.434 | 0.854 |
| AIS C – AIS D | 0.136 | 0.785 | 0.992 |
| Carbon dioxide | | | |
| AIS A – AIS B | -0.581 | 8.22E-02 | 0.287 |
| AIS A – AIS C | 0.298 | 0.251 | 0.642 |
| AIS A – AIS D | 1.635 | 1.29E-03 | **6.00E-03** |
| AIS B – AIS C | 0.879 | 2.06E-02 | 8.68E-02 |
| AIS B – AIS D | 2.216 | 1.28E-04 | **5.33E-04** |
| AIS C – AIS D | 1.337 | 1.29E-02 | 5.67E-02 |
| Neutrophils | | | |
| AIS A – AIS B | -0.153 | 0.851 | 0.997 |
| AIS A – AIS C | -2.315 | 2.64E-04 | **1.37E-03** |
| AIS A – AIS D | -1.563 | 0.205 | 0.565 |
| AIS B – AIS C | -2.162 | 1.96E-02 | 8.33E-02 |
| AIS B – AIS D | -1.41 | 0.316 | 0.734 |
| AIS C – AIS D | 0.752 | 0.565 | 0.935 |
| Lymphocytes | | | |
| AIS A – AIS B | 0.459 | 0.502 | 0.902 |
| AIS A – AIS C | 2.248 | 2.44E-05 | **1.09E-04** |
| AIS A – AIS D | 2.339 | 2.38E-02 | 9.93E-02 |
| AIS B – AIS C | 1.788 | 2.14E-02 | 8.98E-02 |
| AIS B – AIS D | 1.88 | 0.111 | 0.364 |
| AIS C – AIS D | 0.092 | 0.933 | 1 |
| Monocytes | | | |
| AIS A – AIS B | -0.103 | 0.654 | 0.968 |
| AIS A – AIS C | 0.108 | 0.543 | 0.925 |
| AIS A – AIS D | -0.197 | 0.570 | 0.937 |
| AIS B – AIS C | 0.211 | 0.417 | 0.840 |
| AIS B – AIS D | -0.095 | 0.811 | 0.995 |
| AIS C – AIS D | -0.305 | 0.406 | 0.830 |
| Eosinophils | | | |
| AIS A – AIS B | 0.17 | 0.380 | 0.806 |
| AIS A – AIS C | 0.218 | 0.149 | 0.452 |
| AIS A – AIS D | 0.207 | 0.483 | 0.890 |
| AIS B – AIS C | 0.048 | 0.828 | 0.996 |
| AIS B – AIS D | 0.037 | 0.913 | 0.999 |
| AIS C – AIS D | -0.011 | 0.971 | 1 |
| Basophils | | | |
| AIS A – AIS B | -0.061 | 0.104 | 0.346 |
| AIS A – AIS C | 0.004 | 0.878 | 0.999 |
| AIS A – AIS D | 0.077 | 0.178 | 0.514 |
| AIS B – AIS C | 0.065 | 0.124 | 0.396 |
| AIS B – AIS D | 0.138 | 3.39E-02 | 0.136 |
| AIS C – AIS D | 0.072 | 0.230 | 0.610 |
| Glucose | | | |
| AIS A – AIS B | -2.674 | 0.468 | 0.880 |
| AIS A – AIS C | -8.06 | 4.78E-03 | **2.21E-02** |
| AIS A – AIS D | -8.55 | 0.126 | 0.402 |
| AIS B – AIS C | -5.386 | 0.198 | 0.553 |
| AIS B – AIS D | -5.876 | 0.357 | 0.781 |
| AIS C – AIS D | -0.49 | 0.934 | 1 |
| Hematocrit | | | |
| AIS A – AIS B | 1.314 | 9.50E-04 | **4.44E-03** |
| AIS A – AIS C | 1.742 | 1.97E-08 | **7.02E-08** |
| AIS A – AIS D | 3.952 | 8.42E-11 | **2.55E-10** |
| AIS B – AIS C | 0.428 | 0.344 | 0.767 |
| AIS B – AIS D | 2.638 | 1.38E-04 | **6.12E-04** |
| AIS C – AIS D | 2.21 | 6.05E-04 | **3.07E-03** |
| Hemoglobin | | | |
| AIS A – AIS B | 0.437 | 1.35E-03 | **6.60E-03** |
| AIS A – AIS C | 0.61 | 9.74E-09 | **2.93E-08** |
| AIS A – AIS D | 1.392 | 2.52E-11 | **7.45E-11** |
| AIS B – AIS C | 0.173 | 0.264 | 0.662 |
| AIS B – AIS D | 0.955 | 5.63E-05 | **2.62E-04** |
| AIS C – AIS D | 0.782 | 3.98E-04 | **2.16E-03** |
| Potassium | | | |
| AIS A – AIS B | 0.003 | 0.944 | 1 |
| AIS A – AIS C | 0.05 | 6.87E-02 | 0.248 |
| AIS A – AIS D | -0.063 | 0.239 | 0.625 |
| AIS B – AIS C | 0.048 | 0.239 | 0.623 |
| AIS B – AIS D | -0.066 | 0.284 | 0.692 |
| AIS C – AIS D | -0.113 | 4.60E-02 | 0.177 |
| MCH | | | |
| AIS A – AIS B | -0.131 | 0.484 | 0.890 |
| AIS A – AIS C | 0.19 | 0.190 | 0.537 |
| AIS A – AIS D | 0.607 | 3.33E-02 | 0.133 |
| AIS B – AIS C | 0.321 | 0.130 | 0.410 |
| AIS B – AIS D | 0.738 | 2.29E-02 | 0.0957 |
| AIS C – AIS D | 0.417 | 0.167 | 0.492 |
| MCV | | | |
| AIS A – AIS B | -0.505 | 0.508 | 0.906 |
| AIS A – AIS C | 0.983 | 7.70E-02 | 0.276 |
| AIS A – AIS D | 0.089 | 0.922 | 1 |
| AIS B – AIS C | 1.488 | 7.80E-02 | 0.279 |
| AIS B – AIS D | 0.594 | 0.592 | 0.947 |
| AIS C – AIS D | -0.894 | 0.358 | 0.785 |
| Sodium | | | |
| AIS A – AIS B | 0.14 | 0.604 | 0.951 |
| AIS A – AIS C | 0.461 | 2.70E-02 | 0.111 |
| AIS A – AIS D | 1.754 | 1.63E-05 | **6.42E-05** |
| AIS B – AIS C | 0.321 | 0.293 | 0.704 |
| AIS B – AIS D | 1.614 | 5.07E-04 | **2.61E-03** |
| AIS C – AIS D | 1.293 | 2.68E-03 | **1.25E-02** |
| Thrombocytes | | | |
| AIS A – AIS B | -22099.744 | 1.53E-02 | 6.61E-02 |
| AIS A – AIS C | -11293.998 | 0.112 | 0.367 |
| AIS A – AIS D | -18716.165 | 0.180 | 0.517 |
| AIS B – AIS C | 10805.746 | 0.297 | 0.710 |
| AIS B – AIS D | 3383.579 | 0.831 | 0.996 |
| AIS C – AIS D | -7422.167 | 0.615 | 0.955 |
| Erythocytes | | | |
| AIS A – AIS B | 0.16 | 3.84E-04 | **1.91E-03** |
| AIS A – AIS C | 0.178 | 3.80E-07 | **4.45E-06** |
| AIS A – AIS D | 0.365 | 1.16E-07 | **3.95E-07** |
| AIS B – AIS C | 0.019 | 0.716 | 0.982 |
| AIS B – AIS D | 0.205 | 8.77E-03 | **3.94E-02** |
| AIS C – AIS D | 0.187 | 1.05E-02 | **4.65E-02** |
| ASAT | | | |
| AIS A – AIS B | -7.506 | 6.55E-02 | 0.238 |
| AIS A – AIS C | -6.733 | 3.25E-02 | 0.131 |
| AIS A – AIS D | -13.79 | 2.43E-02 | 0.101 |
| AIS B – AIS C | 0.772 | 0.867 | 0.998 |
| AIS B – AIS D | -6.284 | 0.369 | 0.794 |
| AIS C – AIS D | -7.057 | 0.276 | 0.681 |
| ALAT | | | |
| AIS A – AIS B | -5.841 | 0.347 | 0.771 |
| AIS A – AIS C | -10.005 | 3.75E-02 | 0.148 |
| AIS A – AIS D | -24.247 | 9.78E-03 | **4.39E-02** |
| AIS B – AIS C | -4.163 | 0.555 | 0.930 |
| AIS B – AIS D | -18.406 | 8.56E-02 | 0.297 |
| AIS C – AIS D | -14.242 | 0.152 | 0.459 |
| Total serum | | | |
| AIS A – AIS B | 0.074 | 0.208 | 0.572 |
| AIS A – AIS C | 0.254 | 2.23E-08 | **6.86E-08** |
| AIS A – AIS D | 0.358 | 5.55E-05 | **3.27E-04** |
| AIS B – AIS C | 0.18 | 6.71E-03 | **3.08E-02** |
| AIS B – AIS D | 0.285 | 4.95E-03 | **2.35E-02** |
| AIS C – AIS D | 0.104 | 0.267 | 0.667 |
| Triglycerides | | | |
| AIS A – AIS B | -11.168 | 0.115 | 0.374 |
| AIS A – AIS C | -5.503 | 0.317 | 0.734 |
| AIS A – AIS D | 9.608 | 0.373 | 0.798 |
| AIS B – AIS C | 5.665 | 0.481 | 0.888 |
| AIS B – AIS D | 20.776 | 9.04E-02 | 0.31 |
| AIS C – AIS D | 15.111 | 0.185 | 0.528 |
| Leucocytes | | | |
| AIS A – AIS B | -0.13 | 0.730 | 0.985 |
| AIS A – AIS C | -0.405 | 0.169 | 0.495 |
| AIS A – AIS D | -1.205 | 3.67E-02 | 0.145 |
| AIS B – AIS C | -0.275 | 0.521 | 0.913 |
| AIS B – AIS D | -1.076 | 0.101 | 0.338 |
| AIS C – AIS D | -0.801 | 0.190 | 0.538 |

*ASIA impairment scale: A, no sensory or motor function is preserved; B, sensory function is preserved below the level of the injury, but there is no motor function; C, motor function is preserved below the neurological level, and more than half of the key muscles below the neurological level have a muscle grade of <3; D, motor function is preserved below the neurological level, and at least half of the key muscles below the neurological level have a muscle grade of >3.

**Abbreviations**: MCHC: mean corpuscular hemoglobin concentration; MCH: mean corpuscular hemoglobin; MCV: mean corpuscular volume; ASAT: aspartate aminotransferase; ALAT: alanine aminotransferase
