## Supplementary material for "Natural Progression of Routine Laboratory Markers following Spinal Trauma: A Longitudinal, Multi-Cohort Study": Table e-6

| **Supplementary Table 6: Result of pairwise comparisons between the AIS grades on the hematological data from the Murnau study.** | | | |
| --- | --- | --- | --- |
| **Hematological marker** | **Effect** | **p- value** | **Adjusted p- value** |
| Amylase | | | |
| AIS score* (AIS) A – AIS B | -26.704 | 0.013 | 0.061 |
| AIS A – AIS C | -17.292 | 0.104 | 0.356 |
| AIS A – AIS D | -24.272 | 8.64E-04 | **4.42E-03** |
| AIS B – AIS C | 9.412 | 0.482 | 0.892 |
| AIS B – AIS D | 2.432 | 0.824 | 0.996 |
| AIS C – AIS D | -6.98 | 0.512 | 0.910 |
| Alkaline phosphatase | | | |
| AIS A – AIS B | -17.928 | 0.499 | 0.903 |
| AIS A – AIS C | 0.83 | 0.975 | 1 |
| AIS A – AIS D | -6.306 | 0.716 | 0.983 |
| AIS B – AIS C | 18.758 | 0.567 | 0.938 |
| AIS B – AIS D | 11.622 | 0.656 | 0.969 |
| AIS C – AIS D | -7.136 | 0.782 | 0.992 |
| Calcium | | | |
| AIS A – AIS B | 0.02 | 0.400 | 0.829 |
| AIS A – AIS C | 0.014 | 0.558 | 0.934 |
| AIS A – AIS D | 0.082 | 1.34E-07 | **3.87E-07** |
| AIS B – AIS C | -0.006 | 0.830 | 0.996 |
| AIS B – AIS D | 0.062 | 8.39E-03 | **3.97E-02** |
| AIS C – AIS D | 0.068 | 3.00E-03 | **1.53E-02** |
| Cholinesterase | | | |
| AIS A – AIS B | 547.69 | 6.31E-02 | 0.237 |
| AIS A – AIS C | 807.853 | 5.17E-03 | **2.50E-02** |
| AIS A – AIS D | 1468.857 | 7.77E-15 | **2.44E-14** |
| AIS B – AIS C | 260.163 | 0.471 | 0.885 |
| AIS B – AIS D | 921.167 | 1.39E-03 | **6.69E-03** |
| AIS C – AIS D | 661.004 | 1.84E-02 | 8.11E-02 |
| Creatinine | | | |
| AIS A – AIS B | 0.018 | 0.680 | 0.975 |
| AIS A – AIS C | 0.035 | 0.414 | 0.841 |
| AIS A – AIS D | 0.066 | 1.79E-02 | 7.95E-02 |
| AIS B – AIS C | 0.017 | 0.752 | 0.989 |
| AIS B – AIS D | 0.048 | 0.256 | 0.658 |
| AIS C – AIS D | 0.032 | 0.444 | 0.865 |
| CRP | | | |
| AIS A – AIS B | -1.468 | 4.18E-02 | 0.168 |
| AIS A – AIS C | -1.843 | 9.75E-03 | **4.58E-02** |
| AIS A – AIS D | -2.383 | 5.76E-07 | **2.11E-06** |
| AIS B – AIS C | -0.374 | 0.675 | 0.974 |
| AIS B – AIS D | -0.915 | 0.202 | 0.568 |
| AIS C – AIS D | -0.541 | 0.442 | 0.864 |
| Erythrocytes | | | |
| AIS A – AIS B | 0.222 | 4.19E-02 | 0.168 |
| AIS A – AIS C | 0.346 | 1.15E-03 | **5.80E-03** |
| AIS A – AIS D | 0.549 | 4.44E-15 | **1.30E-14** |
| AIS B – AIS C | 0.124 | 0.352 | 0.782 |
| AIS B – AIS D | 0.327 | 2.16E-03 | **1.12E-02** |
| AIS C – AIS D | 0.203 | 4.90E-02 | 0.192 |
| Gamma GT | | | |
| AIS A – AIS B | -34.788 | 0.297 | 0.715 |
| AIS A – AIS C | 43.642 | 0.181 | 0.529 |
| AIS A – AIS D | -57.171 | 7.92E-03 | **3.74E-02** |
| AIS B – AIS C | 78.43 | 5.51E-02 | 0.212 |
| AIS B – AIS D | -22.383 | 0.494 | 0.900 |
| AIS C – AIS D | -100.813 | 1.54E-03 | **7.96E-03** |
| Bilirubin | | | |
| AIS A – AIS B | -0.111 | 0.232 | 0.621 |
| AIS A – AIS C | 0.033 | 0.735 | 0.986 |
| AIS A – AIS D | 0.002 | 0.976 | 1 |
| AIS B – AIS C | 0.143 | 0.221 | 0.602 |
| AIS B – AIS D | 0.112 | 0.217 | 0.595 |
| AIS C – AIS D | -0.031 | 0.746 | 0.988 |
| Total proteins | | | |
| AIS A – AIS B | 0.187 | 0.128 | 0.412 |
| AIS A – AIS C | 0.331 | 5.90E-03 | **2.84E-02** |
| AIS A – AIS D | 0.517 | 8.36E-11 | **3.71E-10** |
| AIS B – AIS C | 0.144 | 0.342 | 0.770 |
| AIS B – AIS D | 0.33 | 6.60E-03 | **3.14E-02** |
| AIS C – AIS D | 0.186 | 0.115 | 0.380 |
| Glucose | | | |
| AIS A – AIS B | 0.346 | 0.480 | 0.891 |
| AIS A – AIS C | 0.484 | 0.336 | 0.763 |
| AIS A – AIS D | 0.472 | 0.151 | 0.465 |
| AIS B – AIS C | 0.138 | 0.825 | 0.996 |
| AIS B – AIS D | 0.125 | 0.798 | 0.994 |
| AIS C – AIS D | -0.013 | 0.980 | 1 |
| Blood urea | | | |
| AIS A – AIS B | 0.214 | 0.570 | 0.939 |
| AIS A – AIS C | 0.378 | 0.308 | 0.730 |
| AIS A – AIS D | -0.18 | 0.465 | 0.881 |
| AIS B – AIS C | 0.164 | 0.724 | 0.984 |
| AIS B – AIS D | -0.394 | 0.290 | 0.706 |
| AIS C – AIS D | -0.558 | 0.124 | 0.405 |
| Hemoglobin | | | |
| AIS A – AIS B | 0.57 | 9.23E-02 | 0.322 |
| AIS A – AIS C | 1.098 | 9.02E-04 | **4.88E-03** |
| AIS A – AIS D | 1.791 | 2.22E-16 | **8.88E-16** |
| AIS B – AIS C | 0.527 | 0.203 | 0.570 |
| AIS B – AIS D | 1.22 | 2.36E-04 | **1.20E-03** |
| AIS C – AIS D | 0.693 | 3.10E-02 | 0.129 |
| Hemoglobin per erythrocyte | | | |
| AIS A – AIS B | -0.117 | 0.731 | 0.985 |
| AIS A – AIS C | 0.109 | 0.741 | 0.987 |
| AIS A – AIS D | 0.268 | 0.213 | 0.587 |
| AIS B – AIS C | 0.226 | 0.587 | 0.946 |
| AIS B – AIS D | 0.385 | 0.250 | 0.649 |
| AIS C – AIS D | 0.159 | 0.618 | 0.958 |
| Hematocrit | | | |
| AIS A – AIS B | 1.858 | 4.68E-02 | 0.185 |
| AIS A – AIS C | 2.943 | 1.26E-03 | **6.26E-03** |
| AIS A – AIS D | 4.887 | 4.44E-16 | **1.33E-15** |
| AIS B – AIS C | 1.086 | 0.342 | 0.771 |
| AIS B – AIS D | 3.03 | 9.42E-04 | **4.65E-03** |
| AIS C – AIS D | 1.944 | 2.85E-02 | 0.120 |
| INR | | | |
| AIS A – AIS B | 0.016 | 0.413 | 0.840 |
| AIS A – AIS C | -0.034 | 6.87E-02 | 0.255 |
| AIS A – AIS D | -0.035 | 4.66E-03 | **2.29E-02** |
| AIS B – AIS C | -0.05 | 3.38E-02 | 0.140 |
| AIS B – AIS D | -0.051 | 6.75E-03 | **3.21E-02** |
| AIS C – AIS D | -0.001 | 0.950 | 1 |
| Potassium | | | |
| AIS A – AIS B | -0.04 | 0.369 | 0.799 |
| AIS A – AIS C | 0.026 | 0.554 | 0.932 |
| AIS A – AIS D | -0.049 | 9.04E-02 | 0.318 |
| AIS B – AIS C | 0.065 | 0.231 | 0.618 |
| AIS B – AIS D | -0.01 | 0.822 | 0.996 |
| AIS C – AIS D | -0.075 | 8.01E-02 | 0.289 |
| Lactate dehydrogenase | | | |
| AIS A – AIS B | 0.394 | 0.978 | 1 |
| AIS A – AIS C | -28.017 | 4.46E-02 | 0.178 |
| AIS A – AIS D | -18.536 | 4.97E-02 | 0.195 |
| AIS B – AIS C | -28.411 | 0.103 | 0.352 |
| AIS B – AIS D | -18.93 | 0.179 | 0.526 |
| AIS C – AIS D | 9.481 | 0.492 | 0.898 |
| Leucocytes | | | |
| AIS A – AIS B | -0.117 | 0.816 | 0.995 |
| AIS A – AIS C | -0.934 | 5.80E-02 | 0.221 |
| AIS A – AIS D | -0.272 | 0.406 | 0.835 |
| AIS B – AIS C | -0.817 | 0.186 | 0.537 |
| AIS B – AIS D | -0.154 | 0.756 | 0.989 |
| AIS C – AIS D | 0.663 | 0.169 | 0.504 |
| Lipase | | | |
| AIS A – AIS B | -14.78 | 0.159 | 0.484 |
| AIS A – AIS C | -3.994 | 0.704 | 0.981 |
| AIS A – AIS D | -14.637 | 4.17E-02 | 0.168 |
| AIS B – AIS C | 10.786 | 0.409 | 0.838 |
| AIS B – AIS D | 0.143 | 0.989 | 1 |
| AIS C – AIS D | -10.643 | 0.310 | 0.733 |
| MCHC | | | |
| AIS A – AIS B | -0.169 | 0.451 | 0.871 |
| AIS A – AIS C | 0.272 | 0.214 | 0.589 |
| AIS A – AIS D | 0.384 | 7.59E-03 | **3.63E-02** |
| AIS B – AIS C | 0.441 | 0.108 | 0.363 |
| AIS B – AIS D | 0.553 | 1.18E-02 | 5.42E-02 |
| AIS C – AIS D | 0.112 | 0.598 | 0.951 |
| MCV | | | |
| AIS A – AIS B | -0.434 | 0.599 | 0.951 |
| AIS A – AIS C | -0.198 | 0.806 | 0.995 |
| AIS A – AIS D | -0.224 | 0.669 | 0.973 |
| AIS B – AIS C | 0.236 | 0.814 | 0.995 |
| AIS B – AIS D | 0.21 | 0.794 | 0.993 |
| AIS C – AIS D | -0.026 | 0.973 | 1 |
| Sodium | | | |
| AIS A – AIS B | 0.925 | 0.153 | 0.470 |
| AIS A – AIS C | 0.391 | 0.538 | 0.924 |
| AIS A – AIS D | 0.852 | 4.27E-02 | 0.171 |
| AIS B – AIS C | -0.534 | 0.502 | 0.904 |
| AIS B – AIS D | -0.073 | 0.909 | 0.999 |
| AIS C – AIS D | 0.461 | 0.457 | 0.875 |
| Prothrombin time | | | |
| AIS A – AIS B | -2.167 | 2.08E-02 | 9.09E-02 |
| AIS A – AIS C | -1.583 | 8.60E-02 | 0.305 |
| AIS A – AIS D | -2.266 | 1.99E-04 | **9.48E-04** |
| AIS B – AIS C | 0.584 | 0.612 | 0.956 |
| AIS B – AIS D | -0.099 | 0.914 | 1 |
| AIS C – AIS D | -0.683 | 0.447 | 0.868 |
| Quick test | | | |
| AIS A – AIS B | 0.874 | 0.750 | 0.988 |
| AIS A – AIS C | 5.317 | 4.79E-02 | 0.188 |
| AIS A – AIS D | 6.892 | 9.76E-05 | **5.97E-04** |
| AIS B – AIS C | 4.443 | 0.186 | 0.538 |
| AIS B – AIS D | 6.018 | 2.52E-02 | 0.108 |
| AIS C – AIS D | 1.575 | 0.547 | 0.928 |
| Thrombocytes | | | |
| AIS A – AIS B | -49.107 | 6.59E-02 | 0.246 |
| AIS A – AIS C | -84.131 | 1.32E-03 | **6.80E-03** |
| AIS A – AIS D | -51.316 | 3.17E-03 | **1.61E-02** |
| AIS B – AIS C | -35.023 | 0.286 | 0.701 |
| AIS B – AIS D | -2.208 | 0.933 | 1 |
| AIS C – AIS D | 32.815 | 0.200 | 0.565 |
| ASAT | | | |
| AIS A – AIS B | -3.736 | 0.644 | 0.966 |
| AIS A – AIS C | -9.117 | 0.259 | 0.662 |
| AIS A – AIS D | -14.638 | 7.62E-03 | **3.61E-02** |
| AIS B – AIS C | -5.381 | 0.594 | 0.949 |
| AIS B – AIS D | -10.903 | 0.181 | 0.529 |
| AIS C – AIS D | -5.522 | 0.493 | 0.899 |
| ALAT | | | |
| AIS A – AIS B | 3.206 | 0.777 | 0.992 |
| AIS A – AIS C | -4.86 | 0.664 | 0.971 |
| AIS A – AIS D | -17.746 | 1.79E-02 | 7.94E-02 |
| AIS B – AIS C | -8.066 | 0.564 | 0.937 |
| AIS B – AIS D | -20.952 | 6.29E-02 | 0.237 |
| AIS C – AIS D | -12.886 | 0.242 | 0.637 |

*ASIA impairment scale: A, no sensory or motor function is preserved; B, sensory function is preserved below the level of the injury, but there is no motor function; C, motor function is preserved below the neurological level, and more than half of the key muscles below the neurological level have a muscle grade of <3; D, motor function is preserved below the neurological level, and at least half of the key muscles below the neurological level have a muscle grade of >3.

**Abbreviations:** CRP: C-reactive protein; Gamma GT: gamma-glutamyl transferase; INR: international normalised ratio; MCHC: mean corpuscular hemoglobin concentration; MCV: mean corpuscular volume; ASAT: aspartate aminotransferase; ALAT: alanine aminotransferase
