## Supplementary material for "Natural Progression of Routine Laboratory Markers following Spinal Trauma: A Longitudinal, Multi-Cohort Study": Table e-7

| **Supplementary Table 7: Comparison of the two data sources.** | | | |
| --- | --- | --- | --- |
| **Hematological marker** | **df** | **Chisq** | **pvalue** |
| Amylase | 1 | 2.66 | 0.103 |
| Alkaline phosphatase | 1 | 0.804 | 0.370 |
| Calcium | 1 | 1.46E+04 | **0.00E+00** |
| Creatinin | 1 | 29.5 | **5.72E-08** |
| Erythrocytes | 1 | 11.2 | **8.08E-04** |
| Bilirubin | 1 | 465 | **3.39E-103** |
| Proteins/Albumin | 1 | 2.09E+03 | **0.00E+00** |
| Glucose | 1 | 1.54E+03 | **0.00E+00** |
| Blood urea | 1 | 346 | **3.10E-77** |
| Hemoglobin | 1 | 28.9 | **7.69E-08** |
| Hematocrit | 1 | 33.2 | **8.29E-09** |
| Potassium | 1 | 34.1 | **5.29E-09** |
| Leucocytes | 1 | 111 | **6.99E-26** |
| MCHC | 1 | 3.77 | 5.23E-02 |
| MCV | 1 | 0.21 | 0.647 |
| Sodium | 1 | 0.612 | 0.434 |
| Prothrombin time | 1 | 160 | **1.39E-36** |
| Thrombocytes | 1 | 421 | **1.65E-93** |
| ASAT | 1 | 19.3 | **1.11E-05** |
| ALAT | 1 | 0.131 | 0.718 |
