## Supplementary material for "Natural Progression of Routine Laboratory Markers following Spinal Trauma: A Longitudinal, Multi-Cohort Study": Figure e-1

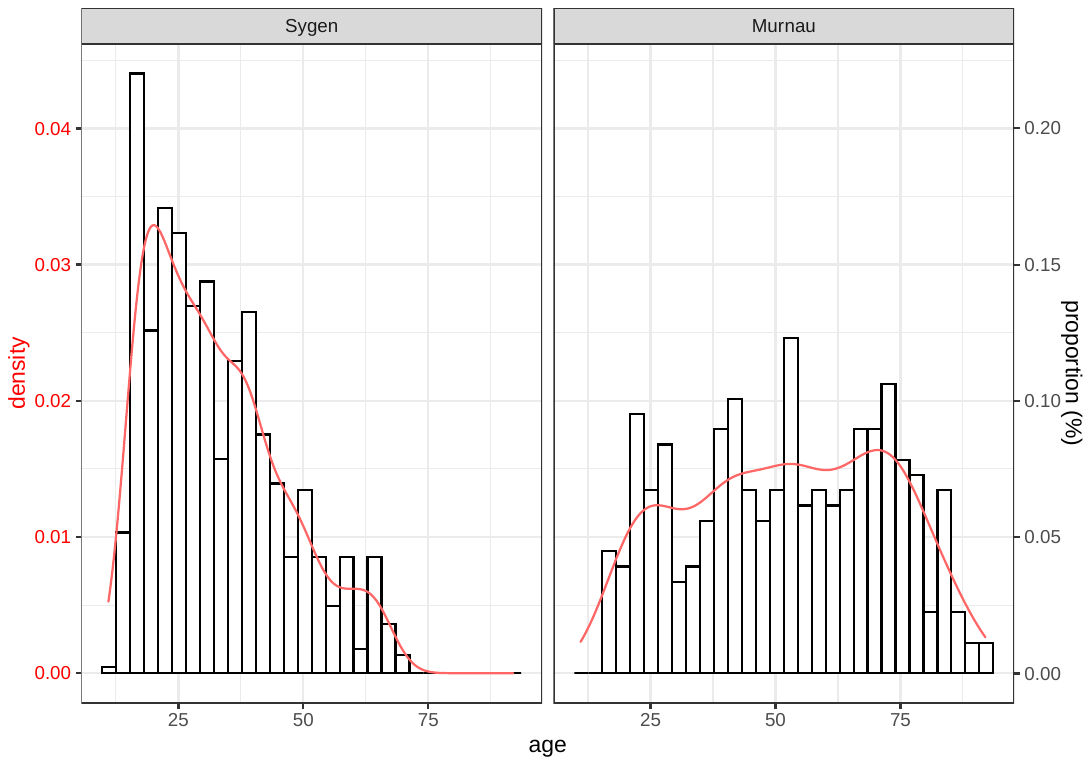

**Figure e-1. Comparison of age at injury distribution of patients enrolled in the Murnau study and Sygen trial.** While mostly young patients were enrolled in the Sygen trial (i.e., left-skewed distribution), the age at injury in the Murnau study was significantly higher (33 ± 14 vs 51 ± 19). The distribution of age at injury in the Murnau approaches a gaussian distribution.
