## Supplementary material for "Natural Progression of Routine Laboratory Markers following Spinal Trauma: A Longitudinal, Multi-Cohort Study": Figure e-5

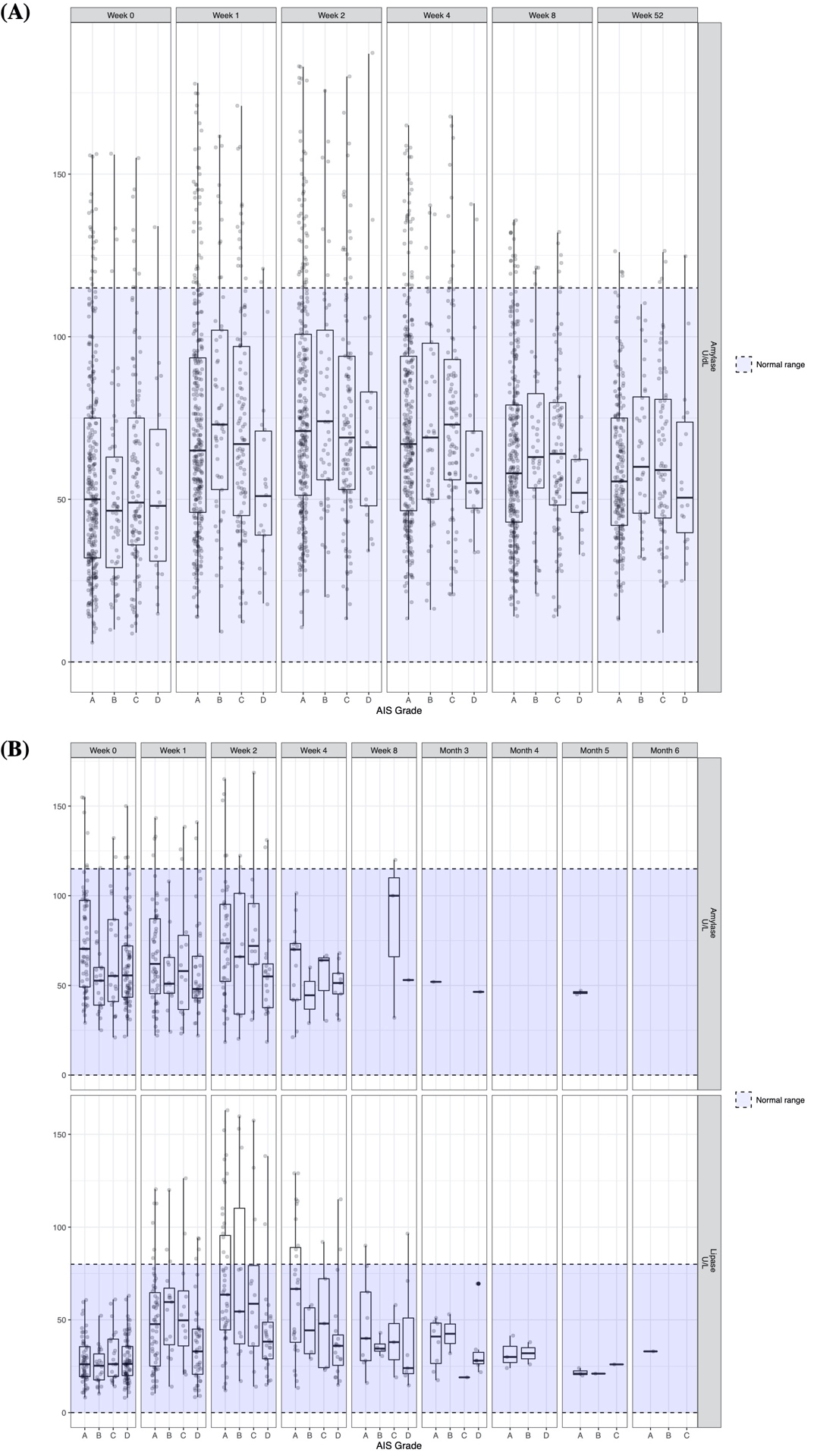

**Figure e-5. Natural progression of blood markers providing information about the pancreas function in patients with spinal cord injury that were enrolled in the Sygen trial (A) and Murnau study (B).**
